# A high throughput blood-based assay for the early detection of pancreatic cancer

**DOI:** 10.1101/2024.01.12.24301220

**Authors:** Jose L. Montoya Mira, Arnaud Quentel, Ranish K. Patel, Dove Keith, Jessica Minnier, Larry David, Sadik C. Esener, Rosalie C. Sears, Charles D. Lopez, Brett C. Sheppard, Utkan Demirci, Melissa H. Wong, Jared M. Fischer

## Abstract

Pancreatic ductal adenocarcinoma (PDAC) remains one of the deadliest cancers due in part to the cancer being diagnosed is at a late stage when effective treatment options are limited. Early detection of PDAC via liquid biopsy would revolutionize survival from the disease. To address the lack of effective non-invasive detection assays for PDAC, we developed a <u>p</u>rotease <u>ac</u>tivity-based assay using a <u>ma</u>gnetic <u>n</u>a<u>n</u>osensor (PAC•MANN). The PAC•MANN assay leverages protease activity in blood to amplify the signal of the target-probe based sensor. An initial screening revealed that the PAC•MANN assay could reliably differentiate patients with PDAC from healthy subjects and patients at high risk of PDAC. Finally, in two cohorts: training (n=145) and blinded validation (n=72), we demonstrated that the PAC•MANN assay had high specificity (86%) and sensitivity (78%) for detection of PDAC compared to healthy subjects. This performance was enhanced when combined with the current standard of care assay, CA19-9 (100% specificity, 84% sensitivity). Our results demonstrate a novel assay that is rapid, high-throughput, and requires low specimen volume, which may not only improve cancer detection but could be useful for monitoring of at-risk patients and could be deployed in low resource settings.

**One sentence summary:** A high-throughput, non-invasive, rapid protease-activated nanosensor identifies pancreatic cancer from a small volume of blood

## INTRODUCTION

Pancreatic ductal adenocarcinoma (PDAC) is frequently not detected until it has progressed to an incurable stage. Associated symptoms are typically non-specific, prolonging the time to diagnoses. Thus, PDAC is on pace to become the second leading cause of cancer-related deaths in the next decade. Effective FDA approved, early detection assays for PDAC detection are non-existent, but are critically needed to transform survival from this deadly disease. The only FDA approved clinical biomarker for PDAC is carbohydrate antigen 19-9 (CA19-9), which is used predominantly for measure of disease burden across treatment and not as an early detection biomarker, due to its low positive predictive value (PPV) in this setting (*1–5*). Currently, our ability to screen the general population as well as monitor patients at high-risk for PDAC is limited by many factors including access, cost, and assay accuracy. Thus, there is an immediate clinical need for the development of biomarker assays that are specific to PDAC, easily accessible, and inexpensive for continual monitoring of cancer risk.

Proteases are an underutilized class of proteins in liquid biopsy-based cancer detection that are directly implicated in tumor progression. Activation of varied proteases is implicated in cancer progression, as they facilitate the degradation of the extracellular matrix and allow cancer cell invasion and dissemination (*6*). In these roles, active proteases are functionally involved in primary cancer progression, dissemination of tumor cells to the blood, and metastatic tumor cell seeding, thus highlighting their potential as a blood-based biomarker for cancer diagnoses (*7, 8*). Active proteases have the potential to increase assay specificity, and their enzymatic activity can also increase assay sensitivity (*9*). The power of developing an assay targeted to tumor-associated proteases is their expansive role in cancer progression, their multi-target cleavage site, and their ability to amplify targets for detection, which offers greater advantages over current conventional protein-based assays for detecting tumor burden. We have developed a novel assay for the unbiased, agnostic measurement of a wide range of proteases that have significant activity across different stages of cancer. Here we show that our assay requires low specimen volume, is high-throughput and, when combined with CA19-9, can detect cancer with 100% specificity and 84% sensitivity. Our assay has the potential to improve pancreatic cancer detection in the general population, amongst high-risk patients and in low-resource and underserved populations.

## RESULTS

### Peptide probe screening for detection of PDAC

We analyzed the effectiveness of an enzymatic activity assay to improve the sensitivity of PDAC detection in peripheral blood. Our initial approach leveraged targets with optimized cleavage sites to capture hundreds of proteases and to expand the number of cleavage sites within each peptide probe. Guided by published assays that had high target sensitivity in small volumes of blood (*10–12*), we optimized a Charge-Changing Protease (CCP) assay that detects a negative-to-positive charge change when the peptide is cleaved, with the resulting products analyzed on a polyacrylamide gel. Uncleaved peptides and serum proteins are positively charged and do not enter the gel, thus enabling a physical separation between the cleaved and uncleaved peptide (**Fig. 1A**). To facilitate unbiased quantification of the cleavage product(s) we developed an automated analysis software (**Fig. S1**). This results in a more accurate and precise evaluation of the total activity signal and increases the dynamic range by two-fold compared to previous quantification methods (p<0.0001, **Fig. S2**). We established the limit of detection to be in the low atto-to femto-moles depending on probe:protease combination (**Fig. 1B)**. This level of detection is nearly 100 times more sensitive than a typical sensitive ELISA for protein expression.

**Figure 1.**
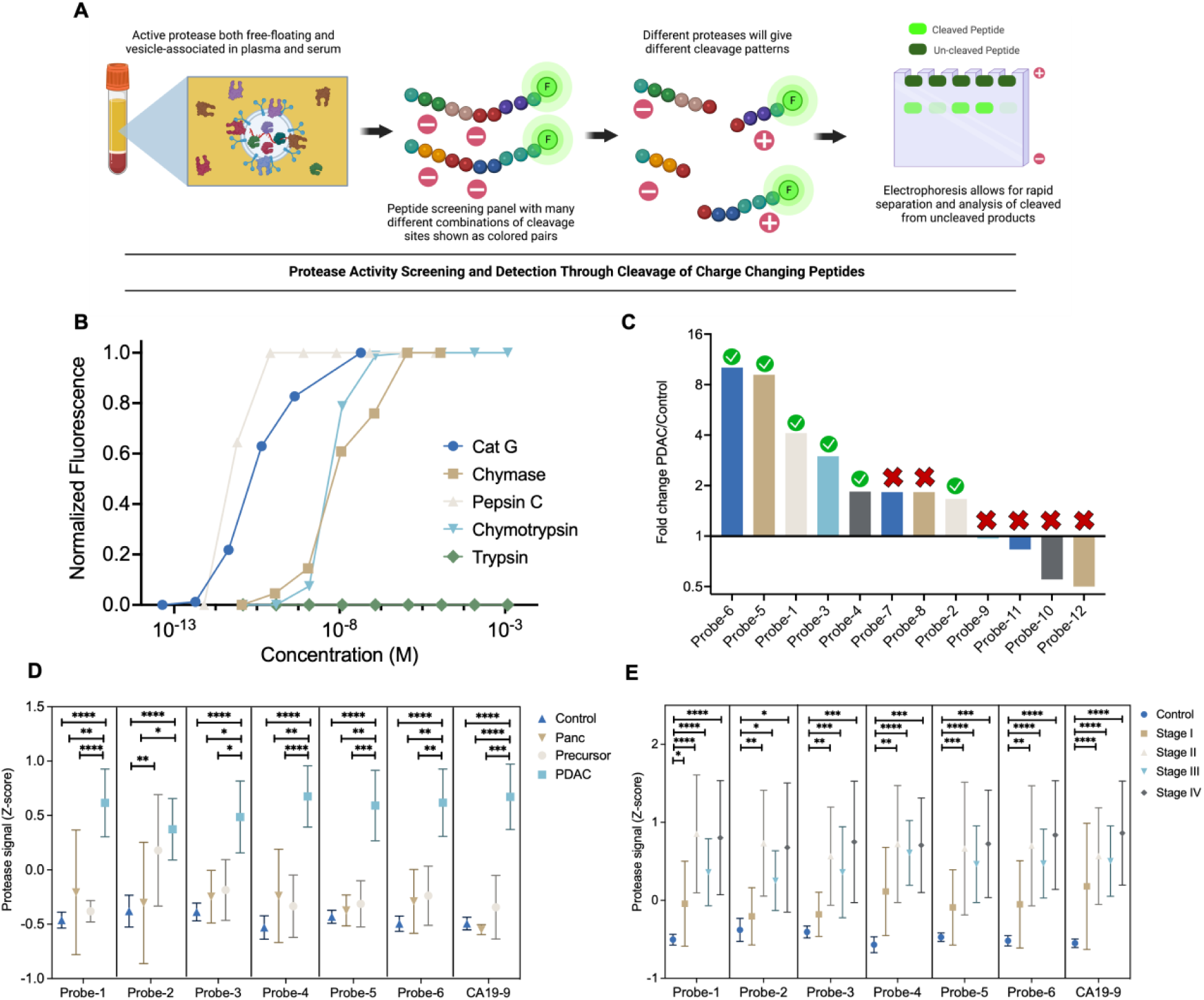
CCP assay to distinguish PDAC from healthy, pancreatitis and pancreatic neoplasias. A) Schematic representation of the approach used to develop protease activity screening panel and detection assay. B) Sensitivity and specificity testing of the charge-changing assay utilizing recombinant enzymes and different concentrations. C) Fold change signal analysis between 6 PDAC and 6 Control. Green check indicates those probes were added to the study while crosses indicated that were not utilized in the main study. D) The Z-score of the protease signal is shown in the graph for every probe with 95% confidence intervals. One-way ANOVA followed by Tukey’s multiple comparisons test for every probe was performed and significance annotated. Control n=67, Pancreatitis (Panc) n=11, Neoplasia n=21, PDAC n=67 E) PDAC patients were subclassified by stage I-IV and their z-score protease signal was plotted for every probe with 95% confidence intervals. One-way ANOVA followed by Tukey’s multiple comparisons test for every probe was performed and significance annotated.

Different proteases target discrete cleavage sites (*13*), thus different peptide probes will have varied specificities in relation to different proteases. To optimize peptide design to serve as targets to a larger number of proteases, we developed a series of peptide probes with a wide range of cleavage sites for many different proteases to enhance efficiency and assay simplicity. To this end, we generated 12 peptide probes that collectively contained 38 unique cleavage sites and 18 shared cleavage sites (p1-p1’) that supported the post-cleavage, change-in-charge readout (**Table S1**). To demonstrate the efficacy of the 12-probe panel, we applied the CCP assay to serum from 6 patients with PDAC (Stage II-IV) and 6 healthy subject controls. Resulting cleavage products were quantified, then by using the average signal from cancer and healthy specimens, we generated a cancer:healthy ratio to identify probes that distinguished PDAC from healthy samples (**Fig. 1C**). We selected 6 probes with the highest cancer:healthy ratios that also harbored the greatest number of unique sites. When combined, the focused 6-probe panel demonstrated strong positive signal in PDAC sera and contained 26 unique cleavage sites and 11 shared cleavage sites. Based upon the *MEROPS* database (*14*), the 37 cleavage sites can be targeted by >100 different proteases, providing the opportunity to capture cancer-associated protease activity that span different phases of pancreatic cancer progression.

### Circulating protease activity distinguishes PDAC from neoplasia and healthy states

To determine the power of the 6 probe-panel to differentiate PDAC from non-cancer, we applied the CCP assay to serum samples from patients with PDAC (n = 67), pancreatic neoplasias (n = 21, including Intraductal Papillary Mucinous Neoplasia [IPMN] and Pancreatic Intraepithelial Neoplasia [PanIN]), chronic pancreatitis (n = 11), and healthy controls (n = 67) (**Table 1 and Table S2**). To individually evaluate each probe for efficacy in differentiating PDAC from healthy controls, we analyzed the Z-scores because each probe had different levels of total fluorescence signal. For all probes, we identified a statistically significant difference between PDAC and healthy controls (p<0.0001, one-way ANOVA followed by Tukey’s multiple comparisons tests; **Fig. 1D**). Across the PDAC disease axis, chronic pancreatitis is an inflammatory disease that harbors an increased risk of PDAC development (*15*), while patients with pancreatic neoplasias (e.g., IPMN, a benign neoplasia (*16*), and PanINs, small lesions with direct link to PDAC (*17, 18*)) also have elevated risk for developing PDAC. In sera from patients with pancreatitis, all 6 probes were detected at levels significantly lower compared to PDAC, but not different from healthy controls. Interestingly, for pancreatic neoplasias, a statistical significance in protease activity was detected in Probe-2 when compared to healthy control sera, while the other 5 Probes distinguished the neoplasias from PDAC (**Fig. 1E**). These data indicate that protease activities are different across the disease axis and suggest that a probe panel can be used to distinguish PDAC from inflammation, neoplasias and healthy controls, while also distinguishing neoplasias from healthy controls.

**Table 1.**
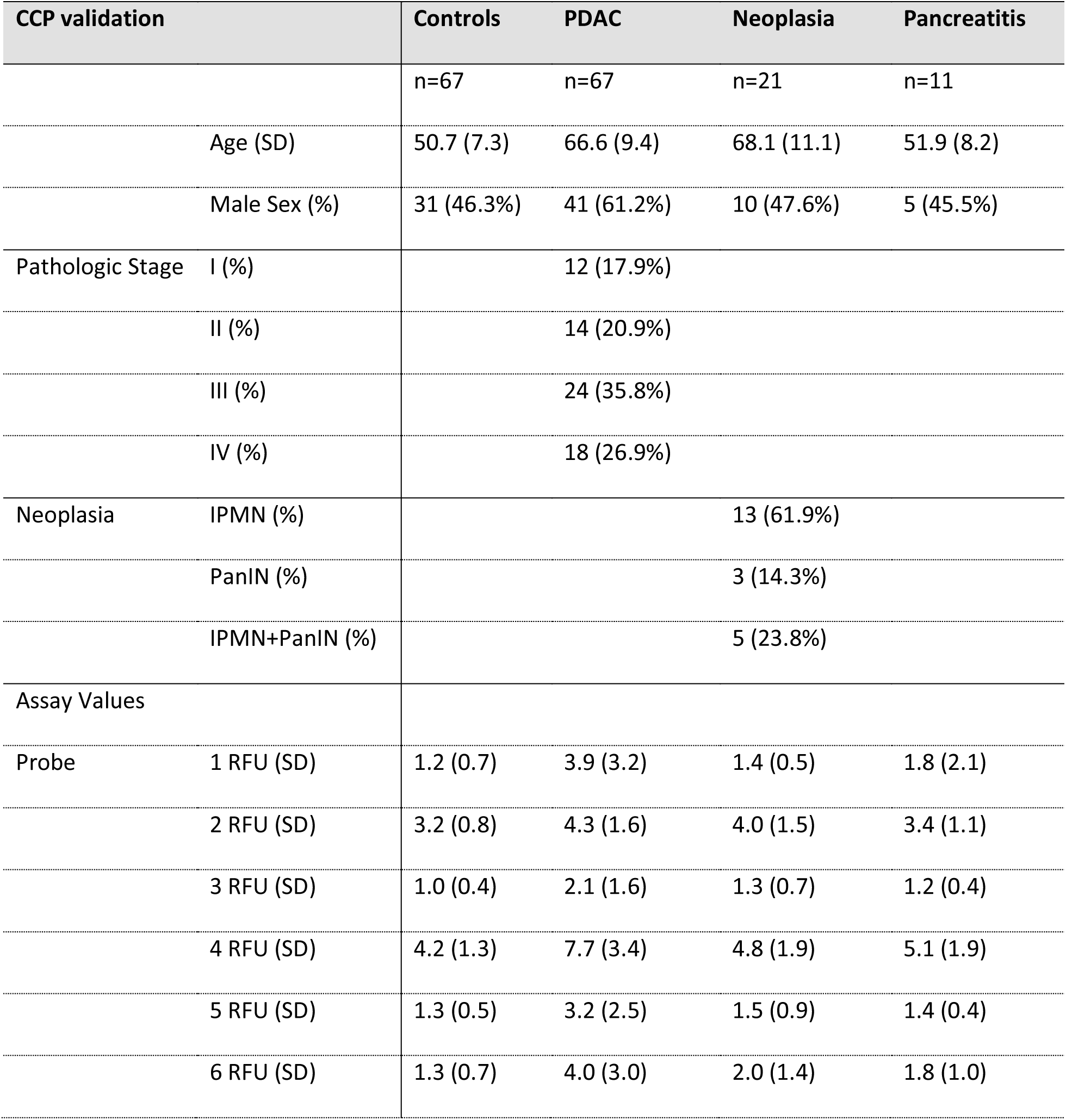
Demographic, clinical characteristics and assay data of patients. Continuous variables are presented as mean (standard deviation [SD]); categorical variables are presented as frequency (%). IPMN – intraductal papillary mucinous neoplasia; PanIN – pancreatic intraepithelial neoplasia.

Finally, for early detection it is important to determine if PDAC can be detected prior to regional dissemination. Thus, PDAC patients were analyzed across pathological stage (I-IV) (**Table 1 and Table S2**) to determine if circulating protease activity differed based on cancer disease burden. Surprisingly, there was no statistically significant difference in protease activity across stage, but protease activity was statistically significant for each stage II, III, and IV patients compared to healthy controls. Interestingly, Probe-1, 3 and 4 showed a significant difference between healthy control patients and early stage (I and II) patients (Multiple t-test with False Discovery Rate correction, **Fig. S3**). While this probe panel effectively differentiates patients with PDAC from healthy subjects, it does not differentiate discrete stages of cancer.

### MMP2 is a key circulating protease in PDAC

While the protease probe panel was created to target a wide number of proteases, we wanted to identify the specific protease(s) that cleave the CCP probes. To identify specific probe cleavage sites, we utilized Liquid Chromatography-Mass Spectrometry (LC-MS) to analyze the cleavage products after incubation with serum from cancer patients (high activity) or healthy subjects (low activity). For example, Probe-1 shows a higher intensity LC-MS peak at site P/F compared to the other possible cleavage sites and the high activity sample is orders of magnitude greater than the low activity sample (**Fig. 2A**). Using this analysis, we calculated the predominant cleavage site for each probe (**Fig. 2B and Fig. S4)**. These results revealed that all 6 peptide probes were predominantly cleaved at different amino acid sites. To identify the possible proteases responsible for probe cleavage, we utilized the ProCleave database that gives cleavage probabilities based upon the sequence from a number of different proteases (*19*). We focused on three protease families that were well represented in the database and upregulated in cancer, MMPs, Cathepsins, and Caspases (*20*). Using the cleavage sites identified by LC-MS, we calculated the number of probes with potential protease activity (**Fig. 2C and Fig. S5)**. We found that MMPs had the highest potential to cleave the greatest number of probes. These results suggested that MMPs, critically important proteases in tumor progression, could be important circulating proteases.

**Figure 2.**
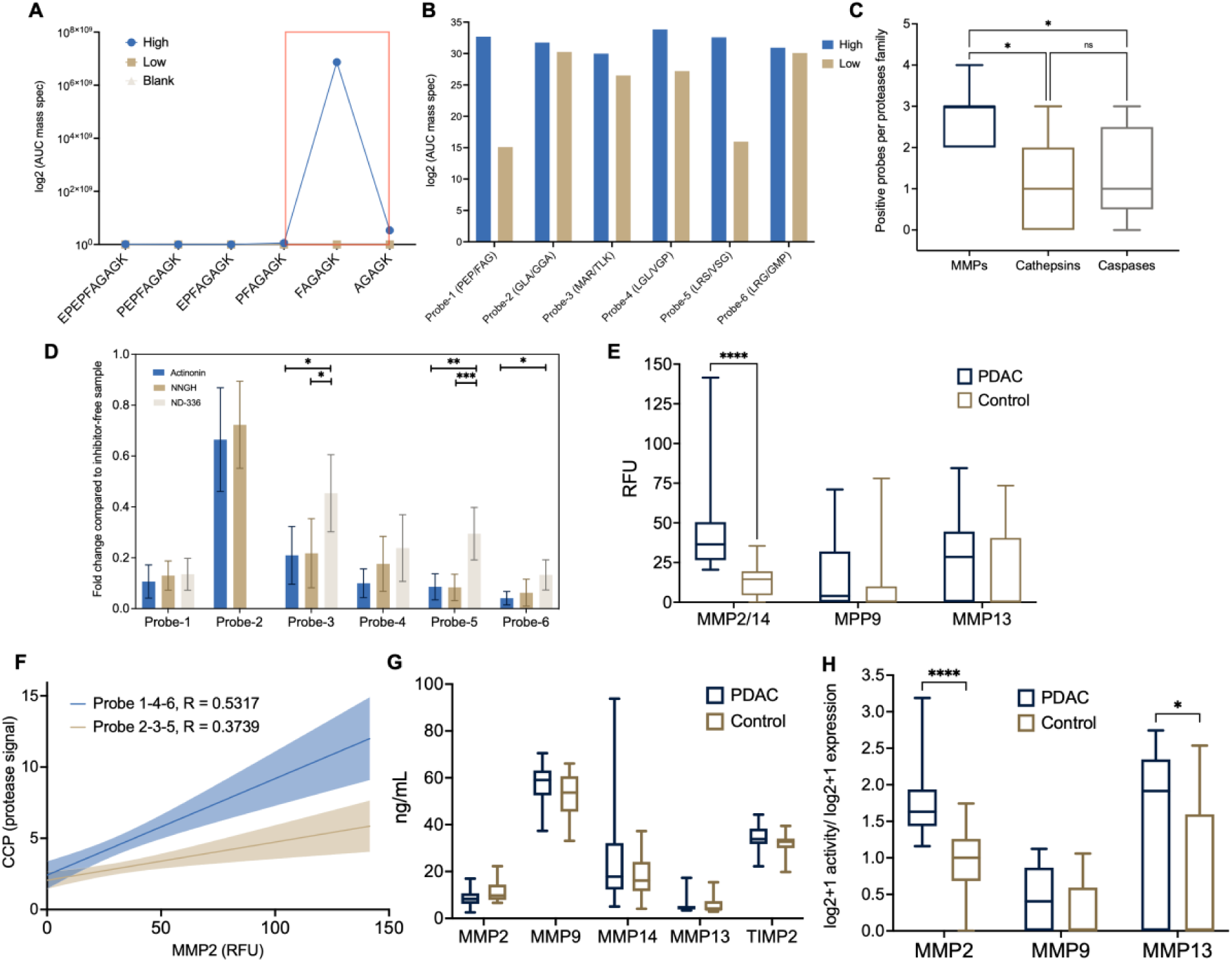
Probe cleavage exploration and active protease identification. A) Mass spectrometry results from isolated probe-1 after incubation with serum to identify cleavage sites. B) Summary of cleavage sites acquired from mass spectrometry results for all probes. C) Number of probes that a single protease can theoretically cleave at the previously identified cleavage site using ProCleave and summarized by protease family. N = 9,7,5 for MMPs, Cathepsins and Caspases respectively, Kruskal-Wallis test with False Discovery Rate correction. D) Identification of main proteases cleaving probes utilizing two MMP pan inhibitors (NNGH and Actinonin) and a specific MMP2/9/14 inhibitor (ND-336). N = 5 for Actinonin with all probes, N = 6 for NNGH with all probes and N= 6 for ND-336 with all probes except probe-2 that was not used. Mean with SD bar plot, One-way ANOVA followed by Tukey’s multiple comparisons test for every probe was performed and significance annotated. Unpaired t-test was performed for probe-2. E) Antibody based activity assay of proteases using a fluorophore-quencher probe mechanism for MMP2, MMP9 and MMP13. Multiple t-test with FDR correction. F) Correlation plot between CCP assay and antibody-specific assay using the average value of probes with higher inhibition level from the specific inhibitor (Probe 1,4,6) and lowest level of inhibition (Probe 2,3,5). G) ELISA of MMP2, MMP9, MMP14, MMP13 and TIMP-2 (Multiple Mann-Whitney test with multiple comparison correction). H) Antibody-specific activity normalized by total protein expression (Multiple Mann-Whitney test with multiple comparison correction).

To further identify key proteases that are enzymatically active in peripheral blood of patients with PDAC, we inhibited discrete MMPs and then examined protease activity in the probe panel. We determined that two pan-MMP inhibitors (Actinonin and NNGH) blocked the majority of enzymatic activity in PDAC serum for 5 of 6 CCP probes (i.e., Probe-1, and 3-6), with a minimal reduction in activity for Probe-2. This finding was consistent with the Procleave database of protease specificity for each probe (**Fig. 2D and Fig. S5**). We next tested inhibitors with greater specificity for MMP-2/9/14 (ND-336). We detected similar inhibition for two probes (i.e., Probe-1 and 4), but only a partial inhibition for three probes (i.e., Probe-3, 5 and 6). These data are again in agreement with the Procleave data as Probe-1 and -4 could be cleaved by a wide range of MMPs (**Fig. S5**). These data strongly implicate the MMP family of proteases having detectible activity in peripheral blood of PDAC patients. Additionally, it is possible that additional protease families are present in peripheral blood of PDAC patients (e.g., Probe-2). For example, Probe-2 is unlikely to be cleaved by MMPs and was the only probe to show differences between healthy controls and patients with pancreatic neoplasias. These results highlight the importance of an agnostic approach for developing a protease-activity-based assay for cancer detection.

To further validate the specific MMPs that are in peripheral blood of patients with PDAC, we used a commercially available assay that first uses antibody-based capture of the target protease and then measures the activity of the captured protease. We tested MMP-2 (with reported minimal cross-reactivity with MMP-14), MMP-9 and MMP-13, and detected significant differences in activity between sera from patients with PDAC and healthy subjects for MMP-2, but not for MMP-9 or MMP-13 (**Fig. 2E**, multiple t-test with FDR correction). To establish the relationship between the specific activity of MMP-2 using the targeted activity assay versus the CCP assay, we performed a correlation matrix for the average of the three probes that displayed highest inhibition levels using a specific inhibitor against MMP-2/9/14 (Probe-1, 4 and 6) and the lowest inhibition (Probe-2, 3 and 5). We determined that CCP probes with higher inhibition correlated better to the MMP2 specific assay (R=0.532) compared to CCP probes with lower inhibition (R=0.374) (**Fig. 2F**, linear regression comparison, p=0.022). Next, we analyzed the total protein levels in sera of MMP-2, MMP-9, MMP-13, MMP-14, and TIMP-2 (an MMP inhibitor protein) and found minimal differences between PDAC patients and healthy subjects (**Fig. 2G**, Mann-Whitney test with multiple comparison correction). Finally, when we normalized enzymatic activity to total protein levels, MMP-2 strongly distinguished PDAC from control specimens (**Fig. 2H**, Mann-Whitney test with multiple comparison correction). These results again implicate active MMP-2 as a key protease disseminated in peripheral blood for the detection of PDAC.

### Highly specific detection of PDAC using a single probe

Since our data suggested that MMP-2 was the main driver of cleavage in our probe set, we analyzed the per-patient correlation of protease activity. As predicted by the biological data, we observe a high correlation between all probes further supporting that a few proteases are driving the majority of cleavage in our probe set (**Fig. 3A**). To further investigate if there were any differences in cancer prediction between each probe individually, as well as the combination of all of them, we performed a logistic regression model for the AUC of the ROC to measure both the sensitivity and specificity of each combination. This analysis revealed that Probe-1 gives a stronger AUC (0.87) compared to any other probe (<0.83) and similar to the combination of all probes (0.86) (**Fig. 3B**). In addition to looking at the ROC, we used backward elimination logistic regression and M-fold cross-validation to test different combinations of probes and determine if there is improved PDAC detection by combining different probes. The combination of all CCP probes had a reduced AUC of 0.82 compared to Probe-1 alone (0.87) (**Fig. S6**), which is mostly due to the multicollinearity of the probes and the specificity of probe-1 for MMP-2 and not other proteases (**Fig. 3A**). These results revealed that the simplest model containing only Probe-1 is best for distinguishing PDAC from healthy controls. Thus moving forward, we concentrated on Probe-1 for PDAC diagnosis. Next, we used cross-validation as a method to test the clinical utility of using Probe-1 for distinguishing PDAC from healthy controls. First, logistic regression was performed on Probe-1 followed by 5-fold cross-validation, in which there is training on 80% of the data, then validation on the remaining 20% of the data over 200 random permutations. This test resulted in an accuracy of 0.79±0.06, AUC of 0.87±0.06, and sensitivity and specificity of 0.68±0.13 and 0.9±0.09, respectively (**Fig. 3C and D**). These results show that a single peptide probe can distinguish PDAC from healthy from a small volume of blood.

**Figure 3.**
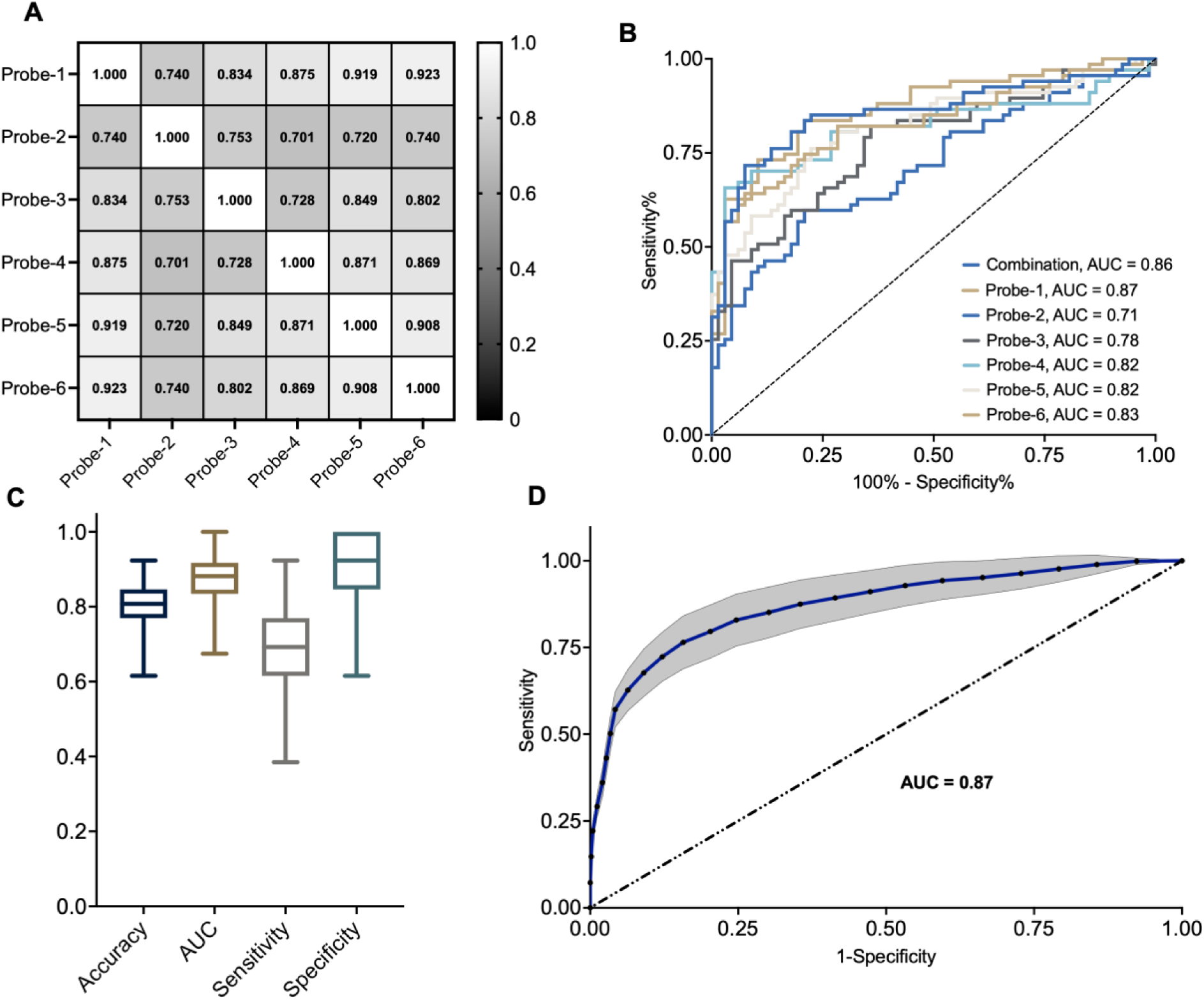
Only a single Probe is needed for PDAC detection. A) Pearson correlation plot between all CCP probes. B) ROC of all probes and their combination. C) Accuracy, AUC of ROC, Sensitivity, and Specificity from cross-validation of Probe-1. D) Cross-validated ROC of Probe-1.

### PAC•MANN assay for clinically relevant PDAC detection

To optimize the clinical translation of the CCP assay for ease of analyses and high-throughput sampling, we developed a <u>p</u>rotease <u>ac</u>tivity-based assay using a <u>ma</u>gnetic <u>n</u>a<u>n</u>osensor (PAC•MANN) that supports a simple fluorescent readout of the cleavage product without the need for gel electrophoresis. For this study, the PAC•MANN assay utilized a D-amino modification of a fluorescently labeled Probe-1 that is covalently attached to the surface of a 50 nm iron oxide nanoparticle, termed PAC•MANN-1. The assay allows for the non-cleaved peptide to be magnetically removed from the solution after incubation with serum, thereby limiting the fluorescent readout to only the cleaved product, while the D-amino acids improve specificity by preventing off-target proteolysis (**Fig. 4A and B**). To demonstrate the benefit of using the nanosensor probe, we analyzed serum from PDAC (n = 27), healthy controls (n = 27) and pancreatic neoplasia samples (n = 38) from subjects with similar average age and sex (**Table S3**). For each sample, we incubated PAC•MANN-1 with 8 μL of serum and detected the cleavage product by fluorescence readout in a 384-well microplate reader. We identified a statistically significance difference between PDAC versus healthy control sera, but no significant difference between healthy and neoplasia sera (**Fig. 4C**). Notably, in comparison with the results from the CCP assay, we detected a strong correlation (R = 0.69), suggesting that we are measuring the same protease activity with both assays (**Fig. 4D**). Finally, to assess the reproducibility and variability of cleavage of PAC•MANN-1, we reanalyzed all PDAC and healthy control sera and observed strong correlation between the technical replicates (R = 0.94); however, the ratio was not 1:1 and there is a need to develop normalization factors between runs (**Fig. 4E**). These results demonstrate the potential for strong clinical performance of PAC•MANN-1 in distinguishing high-risk disease states from PDAC.

**Figure 4.**
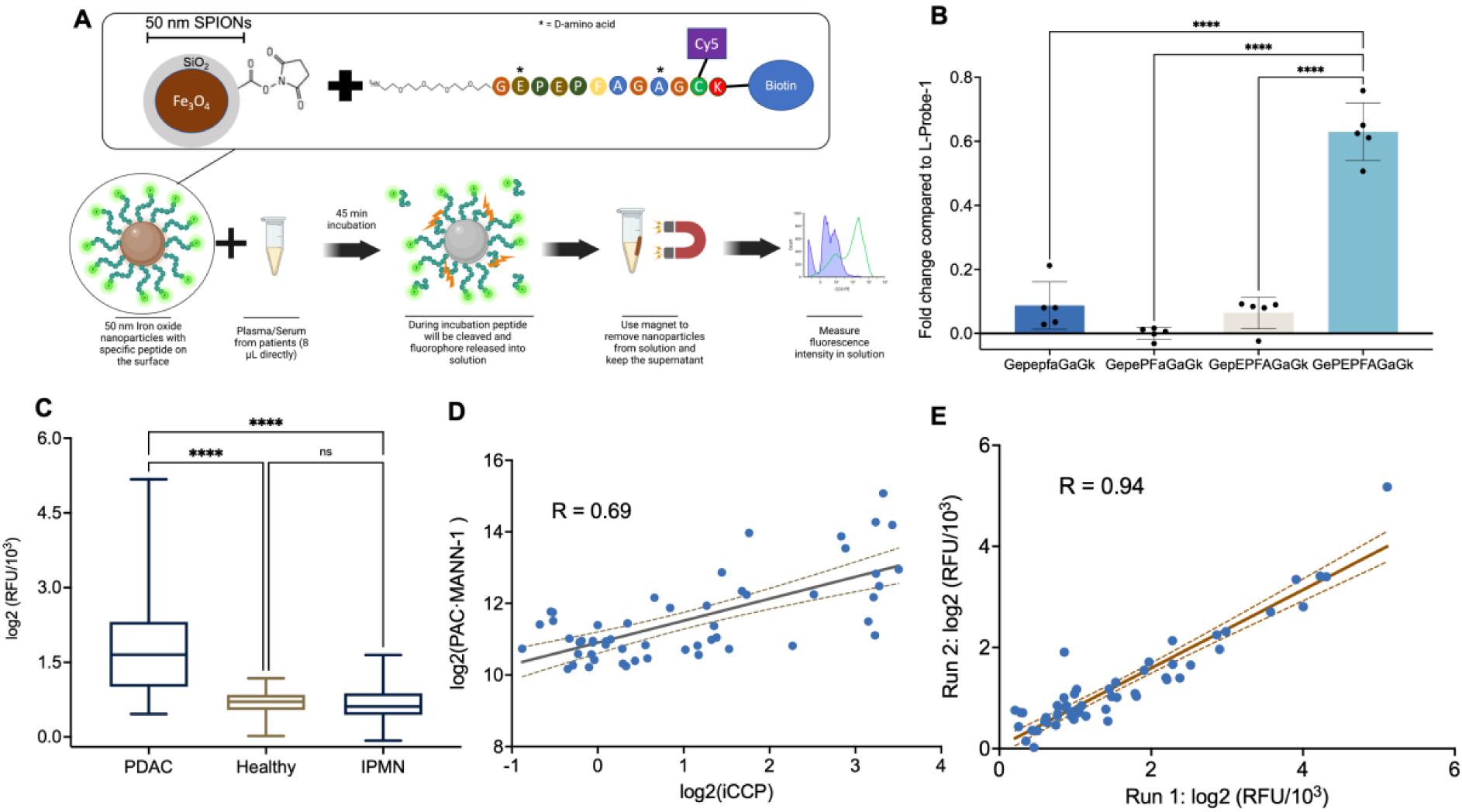
PAC•MANN Assay development for clinical translation. A) Schematic of chemistry used for conjugation. B) Identification of number of amino acids needed for probe to cleave using d-Amino acid modified probes. Mean with SD, One-way ANOVA followed by Tukey comparisons. C) Comparison of 27 PDAC patients, 27 controls and 38 neoplasia (IPMN/PanIN) using PAC•MANN-1 assay (p < 0.0001, Kruskal-Wallis with Dunn’s correction). D) Correlation between CCP assay and PAC•MANN-1 assay using the same 27 PDAC and 27 control patients. E) Correlation between two different batches of PAC•MANN-1 assay.

**Figure 5.**
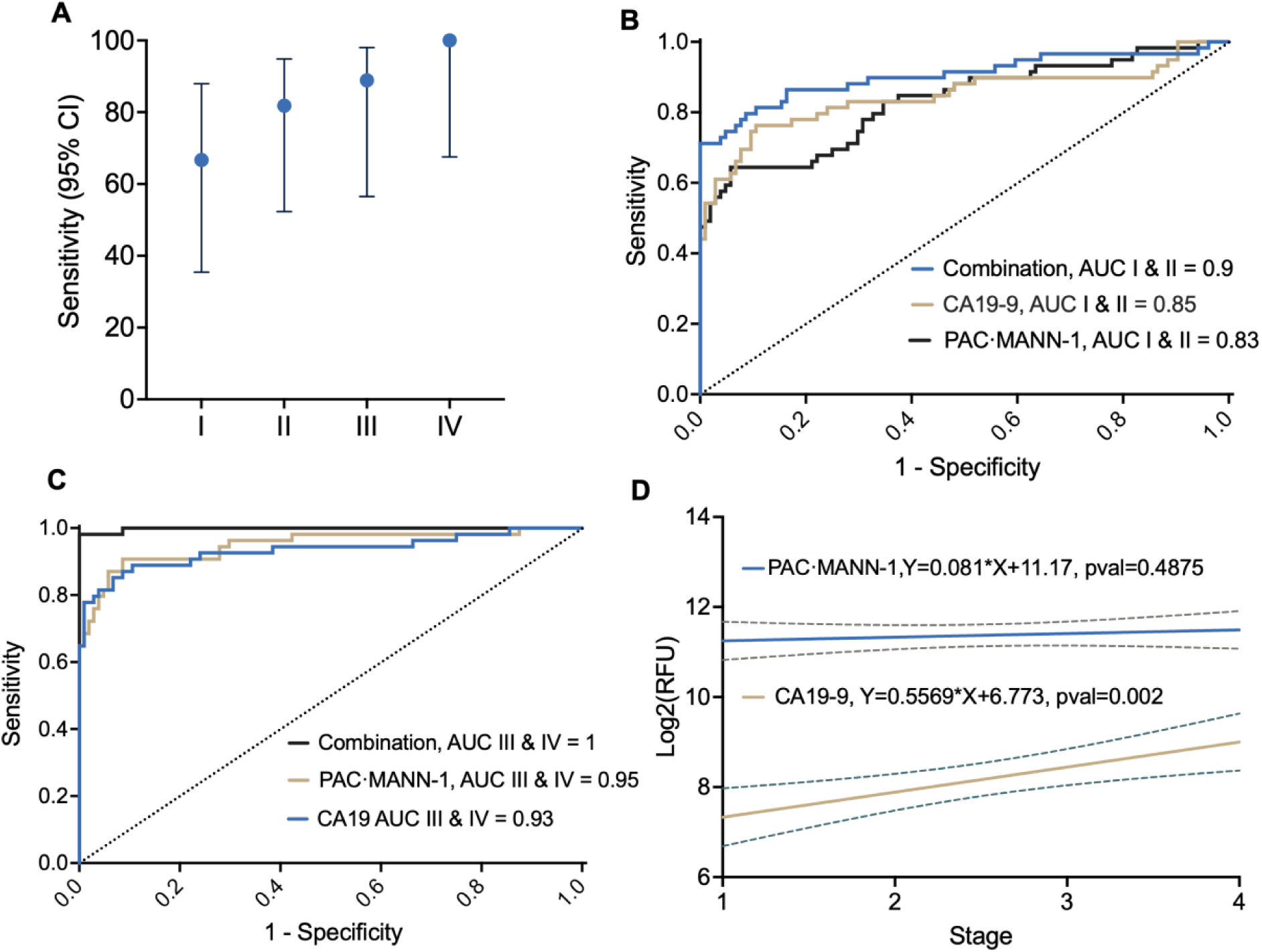
PAC•MANN-1 blinded validation A) Cross-validated sensitivity of the combination of PAC•MANN-1 and CA19-9 (95% CI using Wilson’s method). B) ROC of all combined stage I and II data for the PAC•MANN-1 assay, CA19-9 and the combination. C) ROC of all combined stage III and IV data for the PAC•MANN-1 assay, CA19-9, and the combination. D) Correlation between Stage and CA19-9 and PAC•MANN-1 data.

The initial screening cohort showed that PAC•MANN-1 detected PDAC from healthy and diseased states. To further evaluate PAC•MANN-1, we obtained a larger cohort of patient samples that included an entirely new set of healthy control subjects and 98 additional PDAC patient samples. When added to the previously tested specimens, we evaluated a total of 217 samples (113 PDAC and 104 healthy control) (**Table 2**). To control for batch effects, we ran all 217 samples in a single day. In addition to measuring protease activity, all samples were also analyzed for CA19-9 protein levels. Since the dataset was large enough, we randomly divided the samples by stage into 2/3 for a training data set (n=145) and 1/3 for a blinded validation set (n=72). In the training cohort (69 healthy and 76 PDAC), we found strong statistically significant difference between healthy controls and PDAC samples (p<0.001 Mann Whitney t-test) (**Fig. S7**). CA19-9 also showed strong statistical significance between healthy controls and PDAC (p<0.001 Mann Whitney t-test) (**Fig. S7**). Logistic regression was performed on the training set data and then cutoffs were established for the blinded validation set for the most accurate model, and set specificities of 90%, 96%, and 99%. The data from PAC•MANN-1 were analyzed with and without CA19-9 values in our training and blinded validation cohorts. The AUC of the training set for PAC•MANN-1 alone was 0.86±0.03, for CA19-9 alone it was 0.87±0.03, and for the combination it was 0.93±0.03. These results show that the combination of PAC•MANN-1 plus CA19-9 can distinguish PDAC status compared to healthy individuals.

**Table 2.** Patient Demographic data for the PACMANN-1 validation assay. IQR – InterQuartile Range.

| Characteristic | PDAC Patients<br>(n=113) | n (%) | Healthy<br>Patients<br>(n=104) | n (%) |
| --- | --- | --- | --- | --- |
| Median age at diagnosis: years (IQR) | 69 (62.5-75) |  | 56 (50-64) |  |
| Sex |  |  |  |  |
| Male | 66 | 58.4 | 11 | 10.6 |
| Female | 47 | 41.6 | 93 | 89.4 |
| Race |  |  |  |  |
| White | 102 | 90.3 | 93 | 89.4 |
| Asian | 3 | 2.7 | 0 | 0.0 |
| Unknown | 8 | 7.1 | 11 | 10.6 |
| Ethnicity |  |  |  |  |
| Non-Hispanic or Latino | 107 | 94.7 | 62 | 59.6 |
| Hispanic or Latino | 3 | 2.7 | 1 | 1.0 |
| Unknown | 3 | 2.7 | 41 | 39.4 |
| PAC•MANN-1: RFU (IQR) | 4608 (1383-5103) |  | 1025 (869-1163) |  |
| CA19-9: U/mL (IQR) | 547 (176-769) |  | 58 (18-76) |  |
| Location of Tumor |  |  |  |  |
| Head | 89 | 78.8 |  |  |
| Body | 8 | 7.1 |  |  |
| **Tail** | 8 | 7.1 |  |  |
| **Overlapping** | 8 | 7.1 |  |  |
| **Median Tumor Size:**<br>**mm (IQR)** | 32 (25-42.5) |  |  |  |
| **Pathological Stage** |  |  |  |  |
| **I** | 27 | 23.9 |  |  |
| **II** | 32 | 28.3 |  |  |
| **III** | 29 | 25.7 |  |  |
| **IV** | 25 | 22.1 |  |  |

For the blinded validation, the cutoff established from the training set was used to determine accuracy at each analysis set point (**Table S4**). Overall, we found an 81-85% accuracy for PDAC and healthy patients in our blinded validation cohort using only PAC•MANN-1 or CA19-9 alone. Importantly, combining PAC•MANN-1 and CA19-9 together resulted in an overall accuracy of 92% (66/72 correctly scored). The combination of PAC•MANN-1 plus CA19-9 identified all healthy control samples (100% specificity) and nearly all PDAC samples (84% sensitivity) (**Table S4**). When divided by cancer stage, the combination of PAC•MANN-1 plus CA19-9 set at 100% specificity resulted in sensitivities of 67% for Stage I, 82% for Stage II, 89% for Stage III, and 100% for Stage IV cancers (**Fig. 6A**). These results demonstrated that PAC•MANN-1 combined with CA19-9 increased specificity and sensitivity of PDAC detection from a small volume of blood in a high-throughput and rapid manner.

After analyzing the blinded validation dataset, we combined the training and blinded validation data together into a single dataset of 217 serum samples and reanalyzed the complete dataset. The overall AUC was 0.87 for PAC•MANN-1, 0.89 for CA19-9, and 0.94 for the combination. Importantly, for the combination, at 100% specificity, there was 81% sensitivity. For early detection to be most impactful, it is important to evaluate detection at an early stage (Stages I and II) and demonstrate that an assay can differentiate from normal and early-stage cancer. Therefore, we divided the PDAC samples into early stage (Stage I and II, n=59) and late stage (Stage III and IV, n=54). For early-stage cancers, the AUC was 0.83 for PAC•MANN-1, 0.85 for CA19-9 and 0.9 for the combination (**Fig. 6B**). For late-stage cancers, the AUC was 0.9 for the PAC•MANN-1, 0.93 for CA19-9 and 0.97 for the combination (**Fig. 6C**). In addition to the overall accuracy of the assay, it is also key to understand sensitivity at high specificity, thus for the early-stage cancers at 100% specificity, there was 71% sensitivity. Finally, while PAC•MANN-1 was independent of PDAC Stage (p=0.68 linear regression slope), CA19-9 was positively correlated with PDAC stage (p=0.001 linear regression slope) (**Fig. 6D**). These results indicated that PAC•MANN-1 is highly effective at detecting early-stage cancers and in combination with CA19-9 is highly specific for PDAC, even at early stages.

## DISCUSSION

Development of novel biomarker assays that can efficiently and effectively diagnose lethal cancer at early stages and predict its progression remains an unmet need in PDAC, thus it is a critical aspect of improving survival from cancer. Most cancer detection biomarkers are assayed using hallmarks of apoptosis or rely upon presence of substantial disease burden. Further, some blood-based biomarkers are too rare for reliable sensitivity and reproducibility. Current clinical assays for PDAC rely on detection of the protein CA19-9 and can distinguish high burden cancer from healthy control specimen, but is not sensitive or specific enough to be used as an early detection assay (*21, 22*). In the general, high-risk and underserved populations, a highly accurate screening test that can be performed frequently over time should greatly improve detection and survival.

An ideal early detection, blood-based biomarker would leverage its ability to amplify a small signal or event. Thus, an assay that measures of the activity of a secreted enzyme that is specific to cancer cells compared to normal or neoplastic cells has exciting promise. Cancer-associated proteases secreted into peripheral blood may be relatively rare, but their presence can be leveraged to specifically amplify the cleavage product of a peptide target, thus increasing the readout to robust and detectible levels. Moreover, protease activity is essential for all aspects of cancer progression including invasion, extravasation and intravasation. Herein, we developed a <u>p</u>rotease <u>ac</u>tivity-based assay using a <u>ma</u>gnetic <u>n</u>a<u>n</u>osensor (PAC•MANN) assay that allows for the detection of early-stage pancreatic cancer relative to healthy subjects and patients with neoplasia lesions. We demonstrate that the assay using a single peptide probe (PAC•MANN-1) has high specificity and sensitivity thus reducing the number of false positives and negatives. Further, combining assays for PAC•MANN-1 with CA19-9 improved diagnostic accuracy to a point of having no false positives with only rare false negatives.

One limitation of the current study is the lack of longitudinal specimens, which will be needed to demonstrate efficacy in detecting cancer progression over time. Longitudinal cohorts for early detection are challenging because of the low penetrance of cancers such as PDAC. The number of people that would need to be screened is greater than the cohort that can be feasibly developed. However, PAC•MANN-1 has advantages as the assay is rapid, requires low specimen volume and is low cost, thus could be applied broadly and frequently. Finally, for this assay to have translational application to human health, a robust validation of the assay is needed utilizing thousands of patient samples, which we are currently undertaking. Even with these limitations, we have demonstrated that PAC•MANN-1 may serve as the foundation for a sensitive, low-cost, low volume and scalable early detection of PDAC assay that can be used regularly in high-risk patient populations or the general population with less access to medical tests.

## MATERIALS AND METHODS

### CCP protease conjugation and storage

Peptides were purchased from GenScript and designed with N-terminal acetylation and C-terminal amidation with >98% purity and Trifluoro acetate (TFA) removal. Peptides were dissolved in 100 mM of NaHCO_3_ pH 8.2 to a final concentration of 10 mg/mL. In parallel, we dissolved BODIPY FL NHS Ester (Lumiprobe) in DMSO to a final concentration of 10 mg/mL. Next, we combined equal volumes of both solutions and incubated them at room temperature for 1 hour protected from light. After incubation, the solution was diluted 1:10 in ultrapure water and aliquoted into smaller stock volumes to a final concentration of 500 μg/mL of peptide. Each aliquot was only thawed once to avoid peptide degradation.

### Human Research Participants and Biological Material

We have applied with all ethical guidelines for human research participants. The Institutional Review Board at Oregon Health and Science University provided guidelines for study procedures and protocol was approved (IRB00005169). Informed consent was obtained from all study participants. Blood was collected in serum tubes and allowed to clot for 30 minutes, then the clot was removed by centrifugation at 2,000 x g for 10 minutes. All samples were stored at -80°C. Retrospective samples were used. Sample size of PDAC was based upon obtaining samples that were untreated with similar numbers of each stage. Healthy control samples where there was no known cancer or other reported disease were obtained to match PDAC sample numbers. Pancreatic neoplasia and pancreatitis samples were obtained to give similar numbers to each stage of PDAC, with additional pancreatic neoplasia samples obtained later. Sex was as close to evenly distributed between male and female for the CCP assay and PAC•MANN-1 screening assay, however for the PAC•MANN-1 training and validation the new healthy controls were predominantly obtained from a breast cancer screening cohort.

### CCP protease assay

The general protease assay, unless indicated otherwise, was performed as follows. First 2 μL of 50 mM of calcium chloride is added into a tube or a well of a 96 well plate followed by 4 μL of working stock of peptide and 4 μL of serum. Finally, the solution was mixed and incubated with 150 rpm agitation for 45 minutes at room temperature in the dark. After incubation 8 μL are loaded per well to a 20% acrylamide TBE gel (ThermoFisher) and ran at 250V for 60 minutes with inverted polarity. After the gel has been processed, the gel was imaged using the iBright FL1000 (ThermoFisher) using 488nm channel with 100 ms exposure time unless otherwise specified. Assay was performed non-blinded.

### Quantification for screening

For peptide screening and selection of best targets, we used the previously published methodology for the quantification (*10*). Briefly, ImageJ was used to create a box around the signal of 10x10 pixels and a signal integration was performed and was further used for analysis. Another 10x10 pixel box was created in a region without signal for background subtraction.

### Quantification Software Development

An R script was developed to be able to automatically quantify the fluorescence signal from electrophoretic gels and was specially adapted for each peptide developed in this study. Briefly, the script will take an image where the wells are marked with white lines and will quantify the fluorescence pixel intensity of the signal using an automated integration methodology. The first step is to load the images in tiff file and the script will convert them automatically into a png file. Next, it will compute an image gradient in both the x and y direction and merge them into a single image to be able to detect any borders that are present in the image. After, the script will recognize the y pixel where the wells begin (where the gel start) and the pixels of x where every well starts and ends. After finding the wells, the script will take the average 20 pixels of the center of every well and average them. After averaging them, we will plot a density histogram with the average 20-pixel intensities for all pixels on the y-axis. Next, using experimental data we quantify the length that cleaved probes run into the gel and using that distance in a probe-specific manner and the Simpson rule the script performs a signal integration over 150 pixels. Finally, the integration results are plotted into a histogram where values are indicated and the original gel image will be returned with a rectangle around the areas where the signal was integrated for every well. For every gel processed a pdf is created with a summary of all the plots as well as the final intensity used for further processing.

### Logistic regression model and cross-validation

R was used to assess the diagnostic performance of the different biomarkers. First, an LR model was developed by adding all proteases into the model. After cross validation, the False Positive Rate (FPR) and False Negative Rate (FNR) were both less than 15% (**Figure S6**). In addition, the AUC was greater than 0.8 revealing a strong diagnostic ability of the CCP probe panel (**Figure S6**). Next, backward elimination logistic was performed on proteases to select best-performing targets since individual combined models presented high multicollinearity. After selecting the best-performing protease we used Caret package to assess the performance of the biomarkers. For this purpose, we used an 80/20 split 200-fold cross-validation to asses all possible models and we annotated Sensitivity, Specificity, and AUC of ROC for each fold performed. For each model, we selected the threshold that led to highest accuracy in the training set. Then, this threshold was applied to the held-out portion of the data for testing. The final LR model with nanosensor data was developed and different thresholds were selected and tested in the blinded dataset.

### Liquid chromatography–mass spectrometry (LC–MS)

After incubation of serum samples with each respective peptide, the 8 µL incubation mixtures were frozen before mass spectrometric analysis. Upon thawing, proteins were precipitated by the addition of 24 µL of acetonitrile, samples vortexed, incubated for 5 min, centrifuged at 7,800 x g for 10 min at 20°C, and supernatants removed. A peptide assay was then performed on a portion of the supernatants using a Pierce quantitative colorimetric assay (Thermo Scientific) and indicated that approximately 9 µg of peptide was recovered from each sample. Peptides were then dried by vacuum concentration, dissolved in 5% formic acid and an equal volume of each sample containing approximately 100 ng of peptide was analyzed by liquid chromatography-mass spectrometry (LC-MS). An NCS-3500RS UltiMate RSLCnano UPLC system was used for peptide separation and an Orbitrap Fusion Tribrid instrument with an EasySpray nano source for mass analysis (Thermo Scientific). Peptides were injected onto an Acclaim PepMap 100 μm x 2 cm NanoViper C18, 5 μm trap column on a switching valve. After 5 min of loading and washing, the trap column was switched on-line to a PepMap RSLC C18, 2 μm, 75 μm x 25 cm EasySpray column (Thermo Scientific). Peptides were then separated using a 7.5–30% acetonitrile gradient over 60 min in a mobile phase containing 0.1% formic acid at a 300 nl/min flow rate. Survey mass spectra were acquired over a m/z 175-1600 range in the Orbitrap at a resolution of 120,000 and targeted MS2 scans were performed in the Orbitrap mass analyzer at 7,500 resolution using the calculated m/z values of each peptide, as well as all possible N- and C-terminal fragments calculated using Skyline software (version 21.2.0.425) (MacCoss lab, https://skyline.ms/project/home/software/Skyline/begin.view). Either +1 or +2 charge states for each peptide was used, depending on whether the fragment contained either 1 or 2 basic residues. For each peptide substrate, between 17-35 precursor ions were specified with a loop count set so a survey scan would occur after each group of 10 MS2 scans. Survey scans used a maximum ion time of 50 ms, AGC target of 4 x 105, and were mass corrected using the internal ETD calibrant. Targeted MS2 scans using a quadrupole isolation set at 1.6 m/z, HCD activation at 30% collision energy, maximum injection time of 22 ms, and AGC setting of 5 x104. Results were analyzed using Skyline software, and when possible, the identities of proteolyzed peptide fragments were confirmed by coelution of expected fragment ions in MS2 scans and precursor peptides in survey MS1 scans. Due to their lack of retention on the reverse phase column, single amino acids from the N-terminus were not targeted. However, the BODIPY labeled C-terminal lysine was included (+1 charge state m/z = 420.2377).

### ELISA

ELISA kits were purchased from MyBioSource for MMP-2, MMP-9, MMP-13, MMP-14, TIMP-2 (MBS260339, MBS175780, MBS160467, MBS2516058, MBS355424) and from RayBiotech for CA19-9 (ELH-CA19-9). The protocol was performed as described by the manufacturer. Assays were performed non-blinded.

### Antibody activity assay

Antibody activity assays were purchased from Anaspec and the protocol was performed as described by the manufacturer and the signal was measured after a 1-hour incubation (MMP13 AS-72019, MMP2 AS-72224, MMP9 AS-72017).

### PAC•MANN-1 assay

Absolute Mag™ NHS-Activated Magnetic Particles Conjugation Kit, 50 nm (CD Bioparticles), was used for conjugation, and the protocol was performed as described by the manufacturer with small modifications. Briefly, 50 mg of powder was weighed and added into a 1.5 mL low-bind tube and resuspended with 0.4 mL of activation buffer and vortexing for 15 minutes. Next, a NAP-5 desalting column was used as described by the manufacturer using the activation buffer and after all the beads are inside the column it was moved to a 2 mL low-binding tube and 1 mL of activation buffer added to the column and the beads eluted. Next, 125 nmol of peptide (Nterm-H {PEG4}GePEPFAGaGC(Cy5)K(Biotin)-OH-Cterm) [lower case letters are D-amino acids] (Pepscan) were added to a volume of 250 μL (final) of activation buffer and added to the eluted beads. The solution was vortexed, incubated, and protected from light for 2.5 hours. After incubation 100 μL of quenching buffer was added to the solution and incubated for 30 minutes at room temperature protected from light. After the incubation, 6 washes of the beads were performed using 1 mL storage buffer and waiting progressively shorter time for each wash for the beads to go into the magnet (30 minutes, 30 minutes, 15 minutes, 15 minutes, 5 minutes, and 5 minutes). The last two washes were resuspended 0.5 mL instead of 1 mL. After washing, beads were resuspended in 0.5 mL of storage buffer and stored at 4°C until use.

Just before running the assay, 10 μL from the stock were transferred to a 0.2 mL PCR tube and diluted to 100 μL with storage buffer. Next, 5 washes of the diluted stock were performed using the magnet and 100 μL of storage buffer per wash. After washing the particles were resuspended into 100 μL of storage buffer and the reaction was set up. The reaction was set up in a 0.2 mL PCR tube or 96 well plate low bind. For each reaction, we added 8 μL of beads from the final washed stock, 4 μL of CaCl_2,_ and 8 μL of serum. We pipetted up and down until mixed but with no vortex and placed in a rotator for 45 minutes at room temperature. After incubation, we quickly centrifuged the tubes and added them to the magnet until there was a visible pellet at the back of the tube for around 3 minutes. Finally, we took 15 μL of the supernatant and transferred it to a 384-well plate for fluorescence read-out. We used the TECAN plate reader to analyze Cy5 fluorescence intensity in each condition.

### Statistical Analyses

Measurements were performed on distinct samples. Statistical analyses were performed using GraphPad 9.0 if not indicated otherwise. Normality was assessed using D’Agostino & Pearson test using alpha = 0.05. All statistical analyses were two-sided. Significance nomenclature was used as follows: * = pval < 0.05, ** = pval < 0.01, *** = pval < 0.001, **** = pval < 0.0001. All boxplots above are Min to Max if not indicated otherwise.

### Study Design

Sample size was based upon obtaining >100 samples divided almost evenly between stages. Power analysis was not used for calculating this number. This was a retrospective study, thus there was no stopping of collection, endpoints or inclusion or exclusion criteria. All data were included in the analysis. Each experiment was performed a single time from a blood draw at initial visit and diagnosis of cancer. Our prespecified hypothesis was that protease activity is a biomarker for PDAC alone and in combination with CA19-9. The research subjects were cancer patients, healthy patients, chronic pancreatitis patients, and pancreatic neoplasia patients. This was a controlled laboratory experiment where we measured protease activity, CA19-9 and protein levels. For the screening set, all samples obtained were used. For the training and blinded validation, 2/3 of samples were assigned to the training set and 1/3 were assigned to the validation. Randomization was performed in excel by assigning a random number to each data point, then sorted and divided 2:1. For the training and validation cohorts, all the data was collected at the same time, then the samples were randomized. For the training and blinded validation set, all samples were mixed between cancer and healthy and given a different ID number for blinding during the experiment. For the blinded validation, the samples were assigned a random number and the person analyzing the data and performing experiments was blinded to the sample ID until all analyses were finished.

## Data Availability

All data produced in the present study are available upon reasonable request to the authors

## List of Supplemaentary Materials

**Table S1**. Probe sequences and potential cleavage sites.

**Table S2.** Table of all individual patient demographic and clinical data.

**Table S3.** Patient demographics for the PAC•MANN-1 screening cohort

**Table S4.** Blinded validation results with PAC•MANN-1 assay

**Fig. S1**. CCP quantification software.

**Fig. S2**. Dynamic Range of CCP assay.

**Fig. S3.** Neoplasia versus early stage PDAC.

**Fig. S4.** Cleavage site determination.

**Fig. S5.** Potential protease acting on each probe

**Fig. S6.** Analysis of PDAC and Healthy patients using TSNE and logistic regression.

**Fig. S7.** PAC•MANN-1 and CA19-9 data.

## Authors Contribution

Conceptualization: JLMM, JMF. Methodology: JLMM, MHW, JMF. Software: JLMM. Formal analyses: JLMM, RKP, DK, JM, JMF. Investigation: JLMM, AQ, LD, JMF. Writing original draft: JLMM, JMF. Writing review and editing: JLMM, RKP, MHW, JMF. Visualization: JLMM, RKP, JMF. Supervision: SCE, RCS, CDL, BCS, UD, MHW, JMF. Funding acquisition: SCE, MHW, JMF. **Competing Interests:** UD is a founder of and has an equity interest in: (i) DxNow Inc., a company that is developing microfluidic IVF tools and imaging technologies, (ii) Koek Biotech, a company that is developing microfluidic technologies for clinical solutions, (iii) Levitas Inc., a company focusing on developing microfluidic sorters using magnetic levitation, (iv) Hillel Inc., and Hillal Biotech, companies bringing microfluidic cell phone tools for spermograms to home settings, and (v) Mercury Biosciences, a company developing extracellular vesicle isolation technologies. UD’s interests were viewed and managed in accordance with the conflict-of-interest policies. JLMM and JMF have a patent pending for the PAC•MANN assay. The other authors have no conflict of interest.

