## Supplemental Figures for "A high throughput blood-based assay for the early detection of pancreatic cancer"

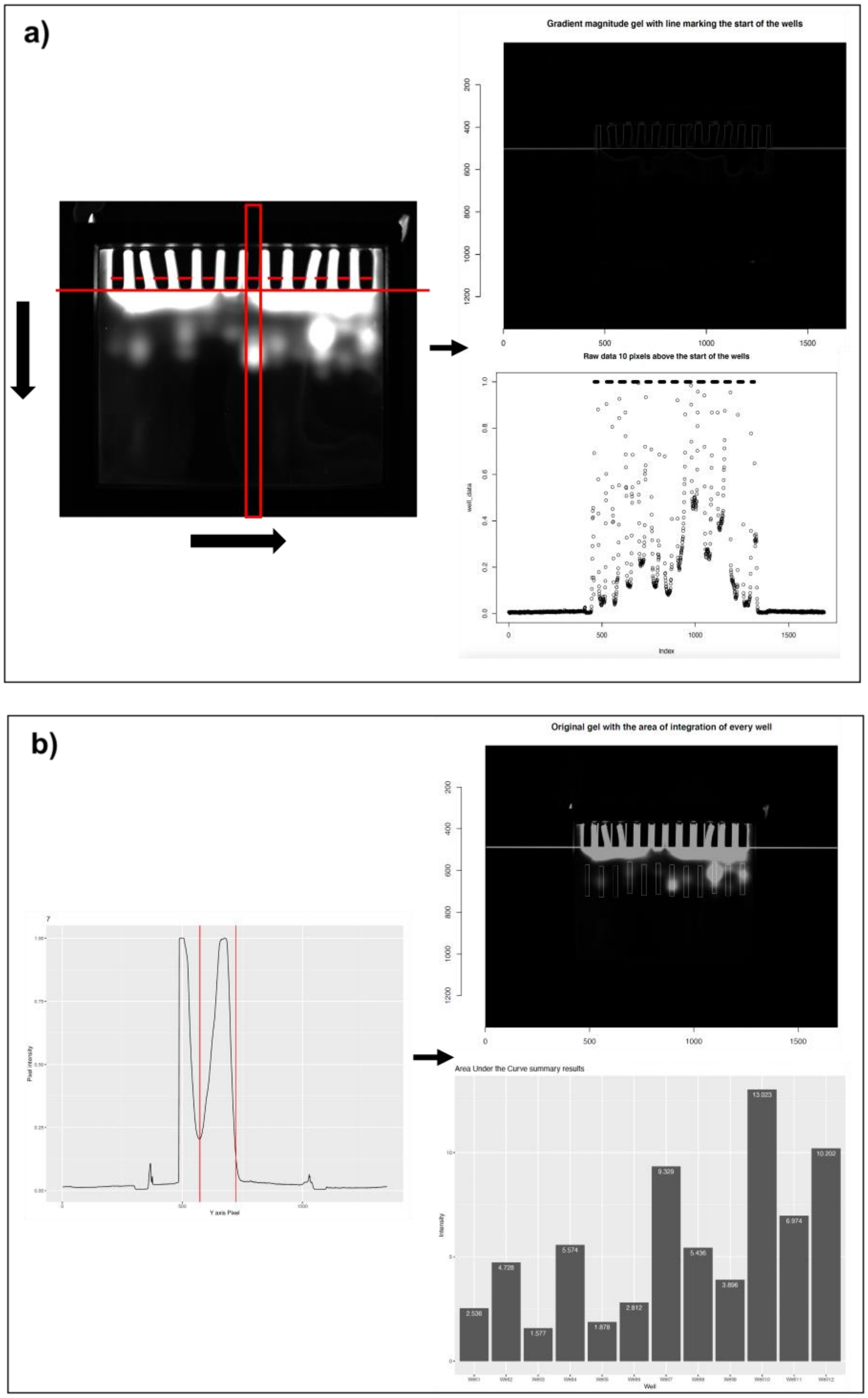


**Fig. S1**. CCP quantification software. Quantification software developed to quantify protease activity using the charge changing peptide technology. A) The software scans through the image to find the teeth of the gel in the x axis as well as the start of the gel inside the well between teeth in the y axis. This enables the identification of all the wells present in the gels. B) For every well, the average signal is plotted across the whole image in the y axis and the signal coming from the peptide is integrated. As a result, an image with white boxes where the integration has been done as well as a bar plot with values per well as created.

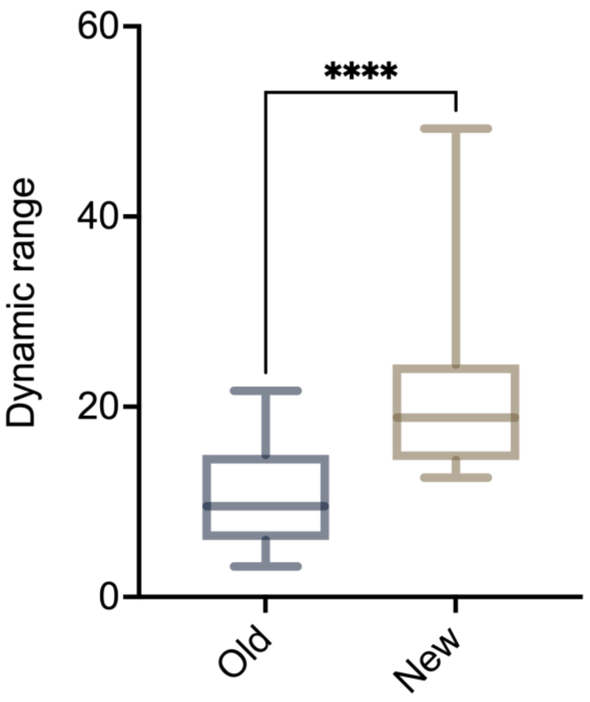


**Fig. S2**. Dynamic Range of CCP assay. Dynamic range comparison between previously published quantification method and newly developed quantification script (n = 34 for old and n = 20 for new, Mann-Whitney test).

| Name | Sequence | Unique cleavage sites | Shared cleavage sites | Unique cleavage sites final assay | Cancer Protease Signal |
|---|---|---|---|---|---|
| *Probe-1* | *GEPEPFAGAGK* | *3* | *3* | *3* | *1.8* |
| *Probe-2* | *DGLAGGAGGK* | *1* | *6* | *1* | *5.4* |
| *Probe-3* | *DGDGMARTLK* | *5* | *2* | *5* | *1.0* |
| *Probe-4* | *DPLGLVGPK* | *1* | *7* | *6* | *4.9* |
| *Probe-5* | *DGDPSLRSVSGK* | *4* | *5* | *5* | *3.0* |
| *Probe-6* | *DGDLRGGMPGSGK* | *5* | *5* | *5* | *4.1* |
| *Probe-7* | *GEIGRLSAGK* | *6* | *2* | | *2.0* |
| *Probe-8* | *DGPAGLVGPK* | *0* | *9* | | *5.8* |
| *Probe-9* | *DLEVLIVLGK* | *4* | *2* | | *0.2* |
| *Probe-10* | *DSSLYSSSGK* | *3* | *4* | | *0.3* |
| *Probe-11* | *DGDGAGYSLPAAGGK* | *3* | *7* | | *0.5* |
| *Probe-12* | *DGDPAELRAGK* | *3* | *5* | | *0.6* |

**Table S1**. Probe sequences and potential cleavage sites.

| Patient ID | Group[$] | Age | Sex | Disease Burden/Pathologic Stage |
|---|---|---|---|---|
| Pancreatitis-1 | | 50-54 | F | Chronic |
| Pancreatitis-2 | | 40-44 | M | Chronic |
| Pancreatitis-3 | | 50-54 | F | Chronic |
| Pancreatitis-4 | | 35-39 | F | Chronic |
| Pancreatitis-5 | | 55-59 | M | Chronic |
| Pancreatitis-6 | | 50-54 | F | Chronic |
| Pancreatitis-7 | | 45-49 | F | Chronic |
| Pancreatitis-8 | | 55-59 | F | Chronic |
| Pancreatitis-9 | | 45-49 | M | Chronic |
| Pancreatitis-10 | | 55-59 | M | Chronic |
| Pancreatitis-11 | | 65-69 | M | Chronic |
| PDAC-1 | Z | 65-69 | M | I |
| PDAC-2 | Z | 70-74 | M | I |
| PDAC-3 | Z | 60-64 | M | I |
| PDAC-4 | | 75-79 | F | I |
| PDAC-5 | P | 60-64 | M | I |
| PDAC-6 | P | 65-69 | M | I |
| PDAC-7 | P | 50-54 | M | I |
| PDAC-8 | P | 65-69 | M | I |
| PDAC-9 | P | 65-69 | F | I |
| PDAC-10 | P | 65-69 | F | I |
| PDAC-11 | P | 65-69 | M | I |

| | | | | |
|---|---|---|---|---|
| PDAC-12 | Z | 70-74 | M | I |
| PDAC-13 | P | 70-74 | M | I |
| PDAC-14 | Z | 65-69 | F | I |
| PDAC-15 | P | 60-64 | M | I |
| PDAC-16 | | 65-69 | F | I |
| PDAC-17 | | 75-79 | M | I |
| PDAC-18 | P | 75-79 | M | I |
| PDAC-19 | Z | 60-64 | M | I |
| PDAC-20 | P | 45-49 | F | I |
| PDAC-21 | P | 55-59 | F | I |
| PDAC-22 | P | 50-54 | F | I |
| PDAC-23 | P | 55-59 | F | I |
| PDAC-24 | P | 70-74 | M | I |
| PDAC-25 | Z | 75-79 | F | I |
| PDAC-26 | P | 65-69 | M | I |
| PDAC-27 | P | 75-79 | F | I |
| PDAC-28 | | 80-84 | M | I |
| PDAC-29 | P | 60-64 | M | I |
| PDAC-30 | Z | 75-79 | F | I |
| PDAC-31 | P | 55-59 | F | I |
| PDAC-32 | Z | 70-74 | M | II |
| PDAC-33 | Z | 80-84 | M | II |
| PDAC-34 | Z | 65-69 | F | II |
| PDAC-35 | Z | 65-69 | F | II |

| | | | | |
|---|---|---|---|---|
| PDAC-36 | Z | 60-64 | M | II |
| PDAC-37 | Z | 65-69 | M | II |
| PDAC-38 | Z | 55-59 | F | II |
| PDAC-39 | | 55-59 | F | II |
| PDAC-40 | Z | 65-69 | M | II |
| PDAC-41 | Z | 60-64 | F | II |
| PDAC-42 | | 60-64 | F | II |
| PDAC-43 | Z | 55-59 | F | II |
| PDAC-44 | Z | 65-69 | F | II |
| PDAC-45 | P | 70-74 | M | II |
| PDAC-46 | P | 35-39 | M | II |
| PDAC-47 | P | 65-69 | F | II |
| PDAC-48 | P | 85-89 | M | II |
| PDAC-49 | P | 55-59 | M | II |
| PDAC-50 | P | 70-74 | M | II |
| PDAC-51 | P | 65-69 | F | II |
| PDAC-52 | P | 70-74 | F | II |
| PDAC-53 | P | 70-74 | F | II |
| PDAC-54 | P | 40-44 | M | II |
| PDAC-55 | P | 65-69 | F | II |
| PDAC-56 | P | 75-79 | M | II |
| PDAC-57 | P | 55-59 | F | II |
| PDAC-58 | P | 55-59 | F | II |
| PDAC-59 | P | 75-79 | M | II |

| | | | | |
|---|---|---|---|---|
| PDAC-60 | P | 60-64 | M | II |
| PDAC-61 | P | 65-69 | F | II |
| PDAC-62 | P | 85-89 | M | II |
| PDAC-63 | Z | 60-64 | M | II |
| PDAC-64 | P | 65-69 | M | II |
| PDAC-65 | P | 75-79 | F | II |
| PDAC-66 | Z | 50-54 | M | III |
| PDAC-67 | Z | 45-49 | F | III |
| PDAC-68 | Z | 45-49 | M | III |
| PDAC-69 | Z | 55-59 | M | III |
| PDAC-70 | Z | 70-74 | F | III |
| PDAC-71 | X | 65-69 | M | III |
| PDAC-72 | Z | 60-64 | F | III |
| PDAC-73 | P | 70-74 | F | III |
| PDAC-74 | Z | 80-84 | F | III |
| PDAC-75 | Z | 70-74 | M | III |
| PDAC-76 | Z | 70-74 | M | III |
| PDAC-77 | Z | 80-84 | M | III |
| PDAC-78 | Z | 70-74 | M | III |
| PDAC-79 | Z | 65-69 | F | III |
| PDAC-80 | Z | 60-64 | F | III |
| PDAC-81 | P | 65-69 | F | III |
| PDAC-82 | P | 50-54 | F | III |
| PDAC-83 | P | 60-64 | M | III |

| | | | | |
|---|---|---|---|---|
| PDAC-84 | P | 70-74 | F | III |
| PDAC-85 | Z | 60-64 | M | III |
| PDAC-86 | Z | 75-79 | M | III |
| PDAC-87 | Z | 75-79 | F | III |
| PDAC-88 | Z | 60-64 | M | III |
| PDAC-89 | Z | 65-69 | M | III |
| PDAC-90 | Z* | 50-54 | M | III |
| PDAC-91 | Z | 65-69 | F | III |
| PDAC-92 | Z | 50-54 | M | III |
| PDAC-93 | Z | 70-74 | M | III |
| PDAC-94 | Z | 65-69 | M | III |
| PDAC-95 | Z | 75-79 | M | IV |
| PDAC-96 | Z | 65-69 | M | IV |
| PDAC-97 | P | 75-79 | F | IV |
| PDAC-98 | Z | 60-64 | M | IV |
| PDAC-99 | P | 55-59 | F | IV |
| PDAC-100 | Z | 40-44 | F | IV |
| PDAC-101 | Z | 75-79 | M | IV |
| PDAC-102 | X | 45-49 | M | IV |
| PDAC-103 | Z | 80-84 | M | IV |
| PDAC-104 | Z | 60-64 | M | IV |
| PDAC-105 | P | 35-39 | M | IV |
| PDAC-106 | P | 40-44 | F | IV |
| PDAC-107 | Z | 75-79 | F | IV |

| | | | | |
|---|---|---|---|---|
| PDAC-108 | Z | 60-64 | M | IV |
| PDAC-109 | Z | 65-69 | F | IV |
| PDAC-110 | Z | 60-64 | F | IV |
| PDAC-111 | Z | 65-69 | M | IV |
| PDAC-112 | Z | 70-74 | M | IV |
| PDAC-113 | Z | 75-79 | M | IV |
| PDAC-114 | Z | 45-49 | F | IV |
| PDAC-115 | | 65-69 | M | IV |
| PDAC-116 | P | 55-59 | M | IV |
| PDAC-117 | P | 55-59 | M | IV |
| PDAC-118 | P | 70-74 | M | IV |
| PDAC-119 | P | 75-79 | M | IV |
| PDAC-120 | P | 65-69 | F | IV |
| PDAC-121 | P | 60-64 | M | IV |
| Neoplasia-1 | X | 65-69 | F | IPMN (low) |
| Neoplasia-2 | X | 70-74 | M | IPMN (low) PanIN 1-3 |
| Neoplasia-3 | X | 55-59 | F | PanIN 1-3 |
| Neoplasia-4 | X | 50-54 | M | IPMN (low) |
| Neoplasia-5 | X | 70-74 | F | IPMN (low) |
| Neoplasia-6 | X | 75-79 | F | IPMN (low) PanIN 1-2 |
| Neoplasia-7 | X | 75-79 | M | IPMN (high) PanIN 2-3 |
| Neoplasia-8 | X | 70-74 | F | PanIN 1, chronic pancreatitis |
| Neoplasia-9 | X | 70-74 | M | IPMN (low) |
| Neoplasia-10 | X | 60-64 | M | IPMN (low) PanIN 1, chronic pancreatitis |

| | | | | |
|---|---|---|---|---|
| Neoplasia-11 | X | 65-69 | M | IPMN (low) PanIN 1 |
| Neoplasia-12 | X | 70-74 | F | IPMN (low) |
| Neoplasia-13 | X | 40-44 | M | IPMN (low) |
| Neoplasia-14 | X | 80-84 | M | IPMN (low) |
| Neoplasia-15 | X | 70-74 | F | IPMN |
| Neoplasia-16 | X | 70-74 | F | IPMN (low) |
| Neoplasia-17 | X | 80-84 | M | IPMN (low) |
| Neoplasia-18 | X | 55-59 | F | PanIN 1, chronic pancreatitis |
| Neoplasia-19 | X | 50-54 | F | IPMN (low) |
| Neoplasia-20 | | 65-69 | F | IPMN |
| Neoplasia-21 | X | 80-84 | M | IPMN |
| Neoplasia-22 | | 75-79 | M | IPMN (high) |
| Neoplasia-23 | X | 70-74 | M | IPMN (low), chronic pancreatitis |
| Neoplasia-24 | X | 70-74 | F | IPMN |
| Neoplasia-25 | X | 80-84 | M | IPMN |
| Neoplasia-26 | X | 70-74 | F | IPMN |
| Neoplasia-27 | X | 70-74 | M | IPMN (low), chronic pancreatitis |
| Neoplasia-28 | X | 65-69 | M | IPMN (high) PanIN 3, chronic pancreatitis |
| Neoplasia-29 | X | 65-69 | F | IPMN (high), chronic pancreatitis |
| Neoplasia-30 | X | 75-79 | F | IPMN (low), chronic pancreatitis |
| Neoplasia-31 | X | 70-74 | F | IPMN (high), chronic pancreatitis |
| Neoplasia-32 | X | 65-69 | M | IPMN (high) |
| Neoplasia-33 | X | 70-74 | M | IPMN (low), chronic pancreatitis |
| Neoplasia-34 | X | 75-79 | M | IPMN (low) |

| | | | | |
|---|---|---|---|---|
| Neoplasia-35 | X | 50-54 | F | IPMN (low), chronic pancreatitis |
| Neoplasia-36 | X | 65-69 | F | IPMN (high) PanIN 1 |
| Neoplasia-37 | X | 60-64 | M | IPMN (low), chronic pancreatitis |
| Neoplasia-38 | X | 65-69 | M | IPMN (low) PanIN 1 |
| Neoplasia-39 | X | 60-64 | F | IPMN (high) |
| Neoplasia-40 | X | 55-59 | F | IPMN (low) |
| Healthy-1 | | 40-44 | M | no known disease |
| Healthy-2 | | 45-49 | M | no known disease |
| Healthy-3 | | 40-44 | F | no known disease |
| Healthy-4 | | 60-64 | F | no known disease |
| Healthy-5 | | 60-64 | M | no known disease |
| Healthy-6 | X | 40-44 | M | no known disease |
| Healthy-7 | X | 40-44 | F | no known disease |
| Healthy-8 | | 55-59 | M | no known disease |
| Healthy-9 | | 40-44 | M | no known disease |
| Healthy-10 | | 55-59 | F | no known disease |
| Healthy-11 | X | 55-59 | F | no known disease |
| Healthy-12 | X | 55-59 | F | no known disease |
| Healthy-13 | X | 45-49 | M | no known disease |
| 60-64 | | 60-64 | F | no known disease |
| Healthy-15 | X | 40-44 | F | no known disease |
| Healthy-16 | | 50-54 | M | no known disease |
| Healthy-17 | X | 45-49 | M | no known disease |
| Healthy-18 | X | 50-54 | M | no known disease |

| | | | | |
|---|---|---|---|---|
| Healthy-19 | X | 50-54 | F | no known disease |
| Healthy-20 | X | 55-59 | M | no known disease |
| 40-44 | X | 40-44 | F | no known disease |
| Healthy-22 | X | 40-44 | F | no known disease |
| Healthy-23 | | 60-64 | F | no known disease |
| Healthy-24 | | 40-44 | M | no known disease |
| Healthy-25 | X | 45-49 | M | no known disease |
| Healthy-26 | | 45-49 | M | no known disease |
| Healthy-27 | | 45-49 | M | no known disease |
| Healthy-28 | | 50-54 | M | no known disease |
| Healthy-29 | X | 40-44 | F | no known disease |
| Healthy-30 | | 45-49 | M | no known disease |
| Healthy-31 | X | 50-54 | F | no known disease |
| Healthy-32 | X | 55-59 | F | no known disease |
| Healthy-33 | | 50-54 | M | no known disease |
| Healthy-34 | X | 40-44 | F | no known disease |
| Healthy-35 | X | 45-49 | M | no known disease |
| Healthy-36 | | 50-54 | M | no known disease |
| Healthy-37 | | 45-49 | F | no known disease |
| Healthy-38 | | 50-54 | F | no known disease |
| Healthy-39 | | 50-54 | F | no known disease |
| Healthy-40 | | 40-44 | M | no known disease |
| Healthy-41 | | 45-49 | F | no known disease |
| Healthy-42 | | 45-49 | M | no known disease |

| | | | | |
|---|---|---|---|---|
| Healthy-43 | X | 50-54 | F | no known disease |
| Healthy-44 | | 55-59 | F | no known disease |
| Healthy-45 | X | 50-54 | F | no known disease |
| Healthy-46 | | 50-54 | M | no known disease |
| Healthy-47 | | 40-44 | M | no known disease |
| Healthy-48 | | 60-64 | F | no known disease |
| Healthy-49 | | 60-64 | M | no known disease |
| Healthy-50 | X | 60-64 | M | no known disease |
| Healthy-51 | | 65-69 | F | no known disease |
| Healthy-52 | | 60-64 | F | no known disease |
| Healthy-53 | | 65-69 | F | no known disease |
| Healthy-54 | | 40-44 | M | no known disease |
| Healthy-55 | X | 45-49 | F | no known disease |
| Healthy-56 | X | 55-59 | M | no known disease |
| Healthy-57 | X | 45-49 | M | no known disease |
| Healthy-58 | X | 40-44 | M | no known disease |
| 45-49 | X | 45-49 | F | no known disease |
| Healthy-60 | | 55-59 | F | no known disease |
| Healthy-61 | | 45-49 | F | no known disease |
| Healthy-62 | | 55-59 | F | no known disease |
| Healthy-63 | | 50-54 | F | no known disease |
| Healthy-64 | | 50-54 | F | no known disease |
| Healthy-65 | Y | 55-59 | M | no known disease |
| Healthy-66 | Z | 50-54 | F | no known disease |

| | | | | |
|---|---|---|---|---|
| Healthy-67 | Z | 45-49 | F | no known disease |
| Healthy-68 | Z | 50-54 | M | no known disease |
| Healthy-69 | P | 60-64 | F | no known disease |
| Healthy-70 | P | 50-54 | M | no known disease |
| Healthy-71 | P | 40-44 | F | no known disease |
| Healthy-72 | P | 70-74 | F | no known disease |
| Healthy-73 | P | 60-64 | F | no known disease |
| Healthy-74 | P | 45-49 | F | no known disease |
| Healthy-75 | P | 70-74 | F | no known disease |
| Healthy-76 | P | 40-44 | F | no known disease |
| Healthy-77 | P | 50-54 | F | no known disease |
| Healthy-78 | P | 50-54 | F | no known disease |
| Healthy-79 | P | 35-39 | F | no known disease |
| Healthy-80 | P | 50-54 | F | no known disease |
| Healthy-81 | P | 45-49 | F | no known disease |
| Healthy-82 | P | 45-49 | F | no known disease |
| Healthy-83 | P | 65-69 | F | no known disease |
| Healthy-84 | P | 70-74 | F | no known disease |
| Healthy-85 | P | 65-69 | F | no known disease |
| Healthy-86 | P | 55-59 | F | no known disease |
| Healthy-87 | P | 60-64 | F | no known disease |
| Healthy-88 | P | 50-54 | F | no known disease |
| Healthy-89 | P | 55-59 | M | no known disease |
| Healthy-90 | P | 60-64 | F | no known disease |

| | | | | |
|---|---|---|---|---|
| Healthy-91 | P | 40-44 | F | no known disease |
| Healthy-92 | P | 55-59 | F | no known disease |
| Healthy-93 | P | 65-69 | F | no known disease |
| Healthy-94 | P | 55-59 | F | no known disease |
| Healthy-95 | P | 55-59 | M | no known disease |
| Healthy-96 | P | 50-54 | F | no known disease |
| Healthy-97 | P | 60-64 | F | no known disease |
| Healthy-98 | P | 55-59 | F | no known disease |
| Healthy-99 | P | 40-44 | F | no known disease |
| Healthy-100 | P | 55-59 | F | no known disease |
| Healthy-101 | P | 65-69 | F | no known disease |
| Healthy-102 | P | 80-84 | F | no known disease |
| Healthy-103 | P | 65-69 | F | no known disease |
| Healthy-104 | P | 50-54 | F | no known disease |
| Healthy-105 | P | 40-44 | F | no known disease |
| Healthy-106 | P | 65-69 | F | no known disease |
| 60-64 | P | 60-64 | F | no known disease |
| Healthy-108 | P | 60-64 | F | no known disease |
| Healthy-109 | P | 50-54 | F | no known disease |
| Healthy-110 | P | 55-59 | F | no known disease |
| Healthy-111 | P | 40-44 | F | no known disease |
| Healthy-112 | P | 50-54 | F | no known disease |
| Healthy-113 | P | 50-54 | F | no known disease |
| Healthy-114 | P | 65-69 | F | no known disease |

| | | | | |
|---|---|---|---|---|
| Healthy-115 | P | 50-54 | F | no known disease |
| Healthy-116 | P | 45-49 | F | no known disease |
| Healthy-117 | P | 65-69 | F | no known disease |
| Healthy-118 | P | 60-64 | F | no known disease |
| Healthy-119 | P | 65-69 | F | no known disease |
| Healthy-120 | P | 40-44 | F | no known disease |
| Healthy-121 | P | 60-64 | F | no known disease |
| Healthy-122 | P | 65-69 | M | no known disease |
| Healthy-123 | P | 60-64 | F | no known disease |
| Healthy-124 | P | 65-69 | F | no known disease |
| Healthy-125 | P | 50-54 | F | no known disease |
| Healthy-126 | P | 70-74 | F | no known disease |
| Healthy-127 | P | 55-59 | F | no known disease |
| 50-54 | P | 50-54 | F | no known disease |
| Healthy-129 | P | 70-74 | F | no known disease |
| Healthy-130 | P | 50-54 | F | no known disease |
| Healthy-131 | P | 55-59 | F | no known disease |
| Healthy-132 | P | 60-64 | F | no known disease |
| Healthy-133 | P | 60-64 | F | no known disease |
| Healthy-134 | P | 55-59 | F | no known disease |
| Healthy-135 | P | 55-59 | F | no known disease |
| Healthy-136 | P | 45-49 | F | no known disease |
| Healthy-137 | P | 45-49 | F | no known disease |
| Healthy-138 | P | 40-44 | F | no known disease |

| | | | | |
|---|---|---|---|---|
| Healthy-139 | P | 70-74 | F | no known disease |
| Healthy-140 | P | 50-54 | F | no known disease |
| Healthy-141 | P | 45-49 | F | no known disease |
| Healthy-142 | P | 30-34 | F | no known disease |
| Healthy-143 | P | 50-54 | F | no known disease |
| Healthy-144 | P | 40-44 | F | no known disease |
| Healthy-145 | P | 60-64 | F | no known disease |
| Healthy-146 | P | 45-49 | F | no known disease |
| Healthy-147 | P | 60-64 | F | no known disease |
| Healthy-148 | P | 50-54 | F | no known disease |
| Healthy-149 | P | 65-69 | F | no known disease |
| Healthy-150 | P | 45-49 | F | no known disease |
| Healthy-151 | P | 60-64 | F | no known disease |
| Healthy-152 | P | 60-64 | F | no known disease |
| Healthy-153 | P | 65-69 | F | no known disease |
| Healthy-154 | P | 50-54 | M | no known disease |
| Healthy-155 | P | 30-34 | F | no known disease |
| Healthy-156 | P | 75-79 | F | no known disease |
| Healthy-157 | P | 50-54 | F | no known disease |
| Healthy-158 | P | 60-64 | F | no known disease |
| Healthy-159 | P | 55-59 | F | no known disease |
| Healthy-160 | P | 35-39 | F | no known disease |
| Healthy-161 | P | 70-74 | F | no known disease |
| Healthy-162 | P | 60-64 | M | no known disease |

| | | | | |
|---|---|---|---|---|
| Healthy-163 | P | 65-69 | M | no known disease |
| Healthy-164 | P | 40-44 | F | no known disease |
| Healthy-165 | P | 55-59 | M | no known disease |
| Healthy-166 | P | 65-69 | M | no known disease |
| Healthy-167 | P | 30-34 | F | no known disease |
| Healthy-168 | P | 65-69 | F | no known disease |
| Healthy-169 | P | 55-59 | M | no known disease |

**Table S2.** Table of all individual patient demographic and clinical data. All blood draws were pre-treatment except where noted. $ (None – patients only included in the CCP assay, X – patients included in both CCP and PAC•MANN-1 screening assay, Z – patients included in both the CCP and PAC•MANN-1 assays, Y – patients only in PACMANN-1 screening assay,  P – patients only included in the PAC•MANN-1 assay. * - patient received neoadjuvant therapy). IPMN is reported as low grade, high grade or unknown

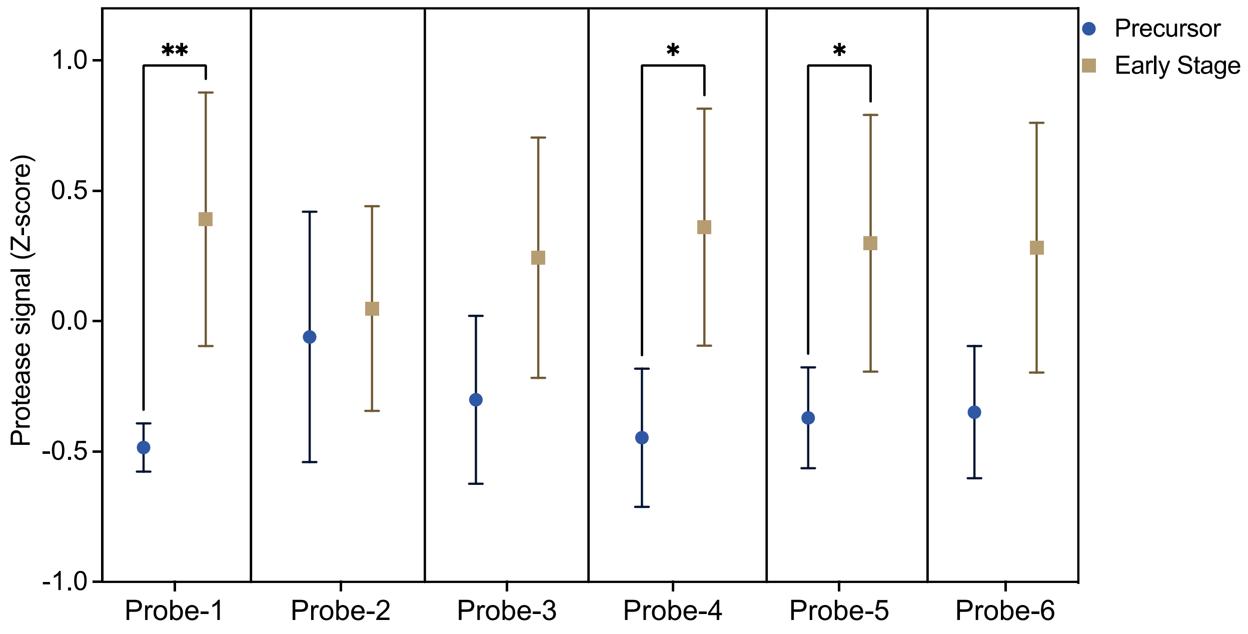


**Fig. S3.** Neoplasia versus early stage PDAC. Analysis of early Stage I and II patients compared to pancreatic neoplasia patients (Multiple t-test with False discovery Rate correction).

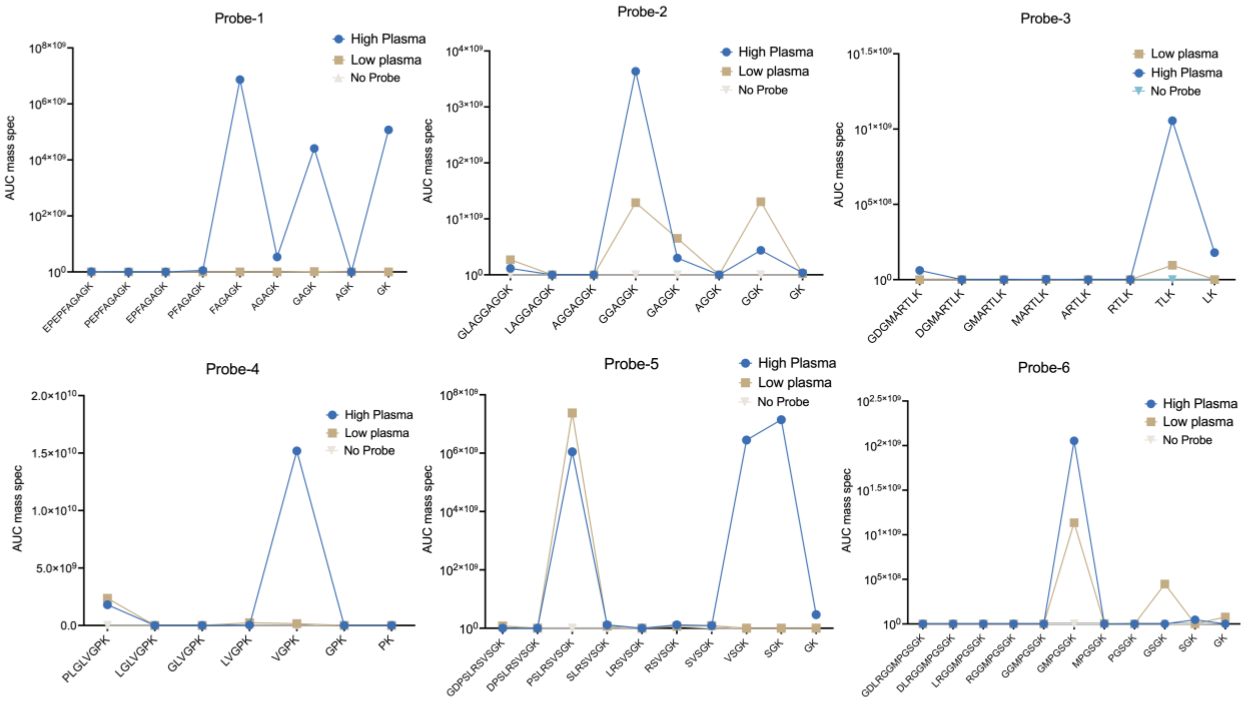


**Fig. S4.** Cleavage site determination. Probe cleavage identification from mass spectrometry results for all probes after incubation with sample with high signal, low signal and sample with high signal but no probe.

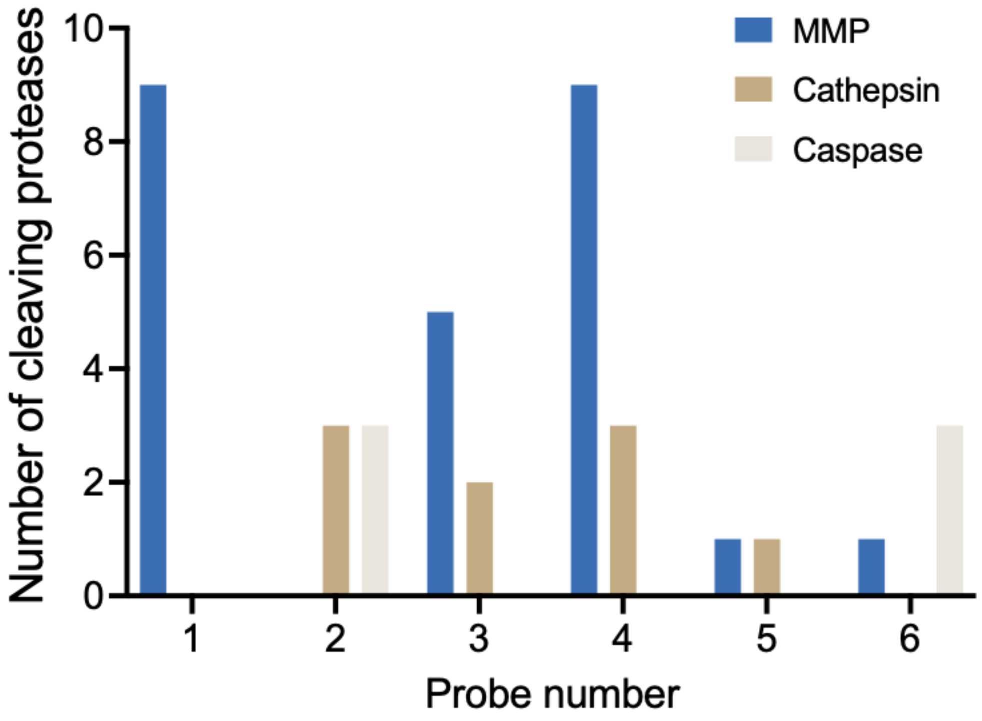


**Fig. S5.** Potential protease acting on each probe. Number of proteases per family that are theoretically able to cleave the probe at the identified cleavage site using ProCleave. MMP=9. Cathepsin=7. Caspase=5.

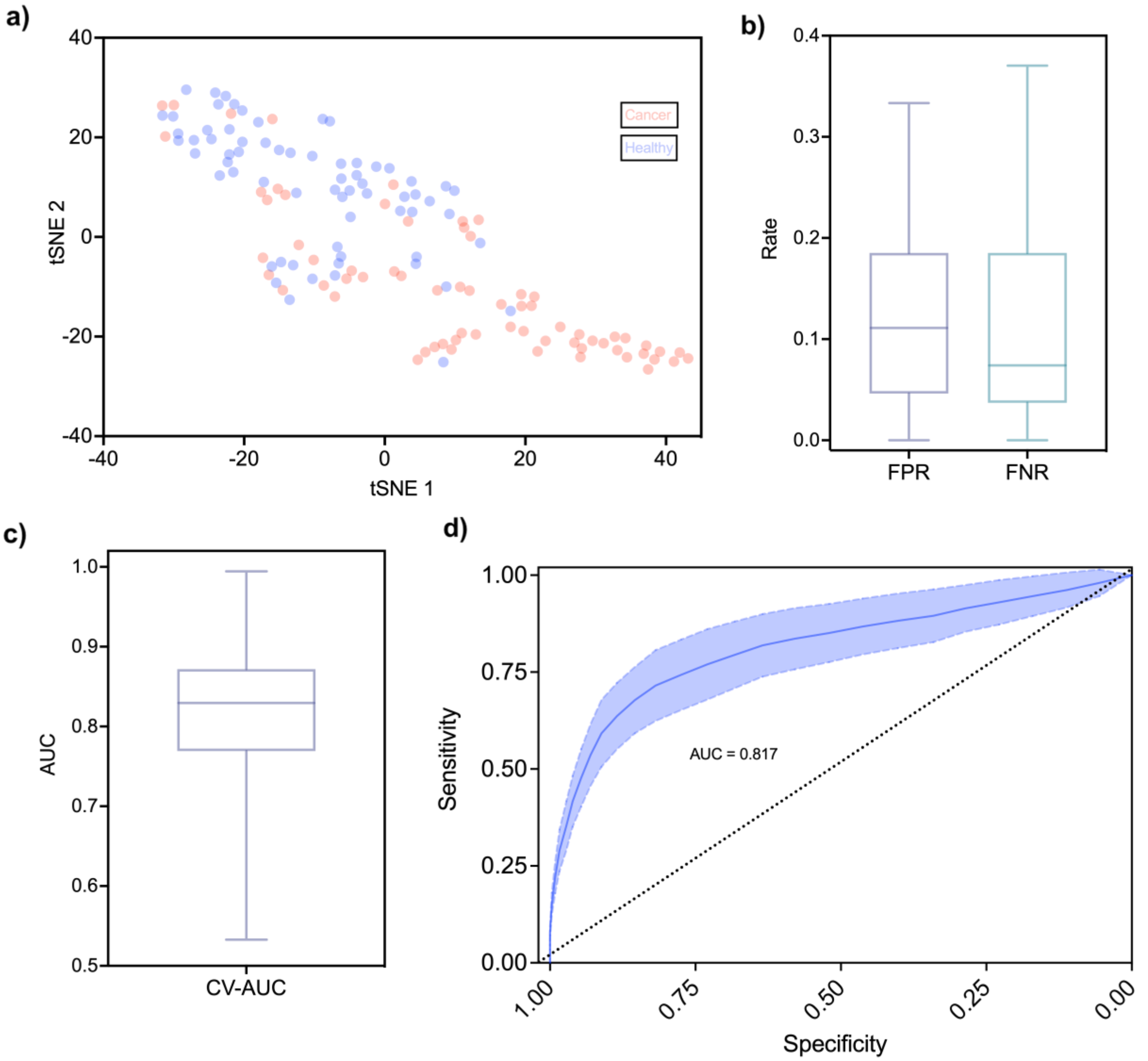


**Fig. S6.** Analysis of PDAC and Healthy patients using TSNE and logistic regression. A) TSNE of protease signal from cancer and healthy is shown. B), C) and D) False positive rate, false negative rate, AUC and ROC from 5-fold cross-validation with 200 random shuffles and 0.5 as probabilistic cut-off.

| **PAC•MANN-1 Screening** | | **Controls** | **PDAC** | **Neoplasia** |
|---|---|---|---|---|
| | | n=27 | n=27 | n=38 |
| | Age (SD) | 49.0 (6.9) | 62.5 (9.1) | 68.6 (9.6) |
| | Male Sex (%) | 12 (44.4%) | 17 (63.0%) | 19 (50%) |
| Stage | I | | 2 (7.4%) | |
| | II | | 7 (25.9%) | |
| | III | | 11 (40.7%) | |
| | IV | | 7 (25.9%) | |
| Disease | IPMN | | | 27 (71.1%) |
| | PanIN | | | 3 (7.9%) |
| | IPMN+PanIN | | | 8 (21.1%) |
| Assay Values | | | | |
| Probe 1 | CCP Z-score (SD) | 1.0 (0.3) | 5.5 (3.4) | |
| | Run 1 RFU (SD) | 1938 (670) | 7281 (7650) | |
| | Run 2 RFU (SD) | 1645 (310) | 5169 (6766) | 1635 (418) |

**Table S3.** Patient demographics for the PAC•MANN-1 screening cohort

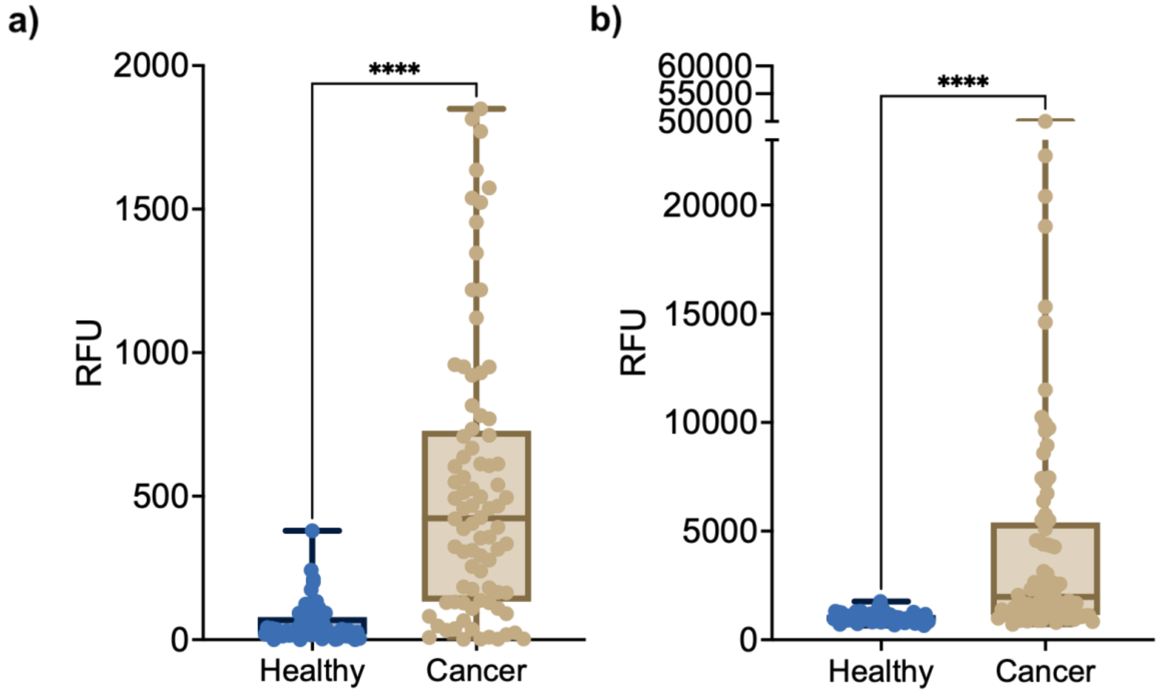


**Fig. S7.** PAC•MANN-1 and CA19-9 data. Training raw data for PAC•MANN-1 (a) and CA19-9 (b) (p<0.001 Mann Whitney t-test).

| Logistic Regression Model | | | | | |
|---|---|---|---|---|---|
| | | 90% cutoff | 96% cutoff | 99% cutoff | Most Accurate |
| **PAC•MANN-1** | | | | | |
| | Specificity | 86 (30/35) | 91 (32/35) | 94 (33/35) | 86 (30/35) |
| **Sensitivity** | I | 78 (7/9) | 78 (7/9) | 67 (6/9) | 78 (7/9) |
| | II | 64 (7/11) | 64 (7/11) | 64 (7/11) | 64 (7/11) |
| | III | 100 (9/9) | 100 (9/9) | 67 (6/9) | 100 (9/9) |
| | IV | 88 (7/8) | 75 (6/8) | 75 (6/8) | 75 (6/8) |
| | all | 81 (30/37) | 78 (29/37) | 67 (25/37) | 78 (29/37) |
| **CA19-9** | | | | | |
| | Specificity | 86 (30/35) | 94 (33/35) | 100 (35/35) | 86 (30/35) |
| **Sensitivity** | I | 78 (7/9) | 56 (5/9) | 56 (5/9) | 78 (7/9) |
| | II | 91 (10/11) | 82 (9/11) | 82 (9/11) | 91 (10/11) |
| | III | 78 (7/9) | 67 (6/9) | 67 (6/9) | 78 (7/9) |
| | IV | 88 (7/8) | 88 (7/8) | 88 (7/8) | 88 (7/8) |
| | all | 84 (31/37) | 73 (27/37) | 73 (27/37) | 84 (31/37) |
| **Combination** | | | | | |
| | Specificity | 83 (29/35) | 91 (32/35) | 97 (34/35) | 100 (35/35) |
| | I | 78 (7/9) | 67 (6/9) | 67 (6/9) | 67 (6/9) |

| | | | | | |
|---|---|---|---|---|---|
| Sensitivity | II | 91 (10/11) | 82 (9/11) | 82 (9/11) | 82 (9/11) |
| | III | 89 (8/9) | 89 (8/9) | 89 (8/9) | 89 (8/9) |
| | IV | 100 (8/8) | 100 (8/8) | 100 (8/8) | 100 (8/8) |
| | all | 89 (33/37) | 84 (31/37) | 84 (31/37) | 84 (31/37) |

**Table S4.** Blinded validation results with PAC•MANN-1 assay
